# Genotype–phenotype correlations and novel molecular insights into the *DHX30*-associated neurodevelopmental disorders

**DOI:** 10.1101/2020.09.24.20196097

**Authors:** Ilaria Mannucci, Nghi D. P. Dang, Hannes Huber, Jaclyn B. Murry, Jeff Abramson, Thorsten Althoff, Siddharth Banka, Gareth Baynam, David Bearden, Ana Beleza, Paul J. Benke, Siren Berland, Tatjana Bierhals, Frederic Bilan, Laurence A. Bindoff, Geir Julius Braathen, Øyvind L. Busk, Jirat Chenbhanich, Jonas Denecke, Luis F. Escobar, Caroline Estes, Julie Fleischer, Daniel Groepper, Charlotte A. Haaxma, Maja Hempel, Yolanda Holler-Managan, Gunnar Houge, Adam Jackson, Laura Kellogg, Boris Keren, Catherine Kiraly-Borri, Cornelia Kraus, Christian Kubisch, Gwenael Le Guyader, Ulf W. Ljungblad, Leslie Manace Brenman, Julian A. Martinez-Agosto, Matthew Might, David T. Miller, Kelly Q. Minks, Billur Moghaddam, Caroline Nava, Stanley F. Nelson, John M. Parant, Trine Prescott, Farrah Rajabi, Hanitra Randrianaivo, Simone F. Reiter, Janneke Schuurs-Hoeijmakers, Perry B. Shieh, Anne Slavotinek, Sarah Smithson, Alexander P.A Stegmann, Kinga Tomczak, Kristian Tveten, Jun Wang, Jordan H. Whitlock, Christiane Zweier, Kirsty McWalter, Jane Juusola, Fabiola Quintero-Rivera, Utz Fischer, Nan Cher Yeo, Hans-Jürgen Kreienkamp, Davor Lessel

**Author notes:** Correspondence should be addressed to N.C.Y, H-J.K or D.L.

## Abstract

**Background:** We aimed to define the clinical and mutational spectrum, and to provide novel molecular insights into *DHX30*-associated neurodevelopmental disorder.

**Methods:** Clinical and genetic data from affected individuals were collected through family support group, GeneMatcher and our network of collaborators. We investigated the impact of novel missense variants with respect to ATPase and helicase activity, stress granule (SG) formation, global translation, and their effect on embryonic development in zebrafish. SG formation was additionally analyzed in CRISPR/Cas9-mediated DHX30-deficient HEK293T and zebrafish models, along with *in vivo* behavioral assays.

**Results:** We identified 25 previously unreported individuals, ten of whom carry novel variants, two of which are recurrent, and provide evidence of gonadal mosaicism in one family. All 19 individuals harboring heterozygous missense variants within helicase core motifs (HCMs) have global developmental delay, intellectual disability, severe speech impairment and gait abnormalities. These variants impair the ATPase and helicase activity of DHX30, trigger SG formation, interfere with global translation, and cause developmental defects in a zebrafish model. Notably, 4 individuals harboring heterozygous variants resulting either in haploinsufficiency or truncated proteins presented with a milder clinical course, similar to an individual bearing a *de novo* mosaic HCM missense variant. Functionally, we established DHX30 as an ATP-dependent RNA helicase and as an evolutionary conserved factor in SG assembly. Based on the clinical course, the variant location and type we establish two distinct clinical subtypes. *DHX30* loss-of-function mutations cause a milder phenotype whereas a severe phenotype is caused by HCM missense mutations that, in addition to the loss of ATPase and helicase activity, lead to a detrimental gain-of function with respect to SG formation. Behavioral characterization of *dhx30* deficient zebrafish revealed altered sleep-wake activity and social interaction, partially resembling the human phenotype.

**Conclusions:** Our study highlights the usefulness of social media in order to define novel Mendelian disorders, and exemplifies how functional analyses accompanied by clinical and genetic findings can define clinically distinct subtypes for ultra-rare disorders. Such approaches require close interdisciplinary collaboration between families/legal representatives of the affected individuals, clinicians, molecular genetics diagnostic laboratories and research laboratories.

## BACKGROUND

RNA helicases (RH) are highly specialized proteins which use ATP hydrolysis for the unwinding of RNA secondary structures and the remodeling of ribonucleoprotein particles (RNPs).[1, 2] RHs are classified into six known superfamilies based on their sequence and structure.[1] Among these, the large helicase superfamily 2 (SF2) contains more than 50 members in humans.[3] These are designated DDX and DHX proteins based on the consensus amino acid sequence DExD or DExH signature in their ATP-binding motif II (Walker B motif).[3] All SF2 RNA helicases are built around a highly conserved helicase core region consisting of two domains that resemble the bacterial recombination protein recombinase A (referred to as RecA-1 and RecA-2). Within these two core helicase domains, eight highly conserved sequence elements, helicase core motifs (HCMs) play a role in either RNA binding, or ATP binding and hydrolysis. The roles of SF2 RNA helicases include regulation of splicing, nuclear mRNA export, translation, transcription, facilitation of mRNA decay, microRNA processing, and cytoplasmic transport and storage of RNAs.[1] Thus far, many of the RHs have been studied in various cancers revealing the role of translation in carcinogenesis,[4] and serve as potential biomarkers for diagnosis and prognosis, and novel drug targets.[5] The importance and functional relevance of certain SF2 RHs in human neurodevelopment is demonstrated by the identification of pathogenic germline variants in *DDX3X,[6] DDX6,[7] DHX30[8]* and *DDX59*[9] in individuals with neurodevelopmental disorders. Additionally, a paralog-based study implicated a role for *DHX16, DHX34, DHX37* and *DDX54*, in human neurodevelopmental disorders and suggested that *DHX8, DDX47* and *DHX58* may also be neurodevelopmental genes.[10]

Previously, we reported 12 unrelated individuals with global developmental delay (GDD), intellectual disability (ID) accompanied by severe speech impairment and gait abnormalities, harboring one of six different *de novo* missense variants located within highly conserved HCMs of *DHX30*.[8] Moreover, a recent study reported gonadal mosaicism in two brothers carrying a *de novo* missense variant, p.(Ser737Phe), which resides within a HCM.[11] Here, we performed clinical, genetic and functional analyses to provide further understanding of *DHX30*-related neurodevelopmental disorders through the identification of 25 previously unreported individuals. This systematic clinical and research approach, partially facilitated through social media, establishes novel genotype-phenotype correlations based on *in-depth* functional analyses accompanied by clinical and genetic findings.

## MATERIALS AND METHODS

### Human subjects and genetic analyses

The study was performed in accordance with protocols approved by the respective ethics committees of the institutions involved in this study (approval number by the Ethics Committee of the Hamburg Chamber of Physicians: PV 3802 and the UCLA IRB: 11-001087). Next-generation sequencing based analyses were performed in various independent research or diagnostic laboratories worldwide, using standard approaches described in the supplementary data (more details are available upon request). Most individuals were enrolled in the present study through the “*DHX30 family support group”* on Facebook: https://www.facebook.com/groups/1808373282809332. In such a case the families/legal representatives were asked to provide the contact details of attending physicians in order to obtain objective and accurate clinical and genetic data. Others presented in the University Medical Center Eppendorf, Hamburg, Germany, or were recruited through GeneMatcher [12] and our network of collaborators. For all individuals, clinical data and information on genetic testing were uniformly obtained from attending physicians using a structured clinical table (Table S1) and clinical summary (Supplementary data).

### Cell culture and *in-vitro* assays

Human embryonic kidney 293T (HEK 293T) cells and human bone osteosarcoma epithelial (U2OS) cells were grown under standard condition as described previously.[8] DXH30 expression vectors based on pEGFP-C3 (leading to an N-terminal GFP-tag) and pEGFP-N2 (for expression of the mitochondrial form of DHX30 with a C-terminal GFP-tag) have been described. Newly identified missense variants were introduced into both vectors using Quick-Change II site directed mutagenesis kit (Agilent, Waldbronn, Germany.[8] Transient transfection of HEK293T and U2OS cells, ATPase assay (in HEK293T), and immunocytochemistry and puromycin incorporation assay (in U2OS) were performed as previously described.[8]

### Helicase assay

6xHis-SUMO-DHX30 wild-type and mutant proteins were expressed in the E. coli BL21 (DE3) pLysSpRARE cells (Novagen, Germany). Proteins were purified from lysates using Ni-NTA beads (Qiagen, Germany) as previously described.[13] To test the RNA unwinding activity of DHX30, a [^32^P]-labeled RNA duplex was synthesized using the T7 RNA polymerase from a linearized DNA template designed by Tseng-Rogenski and Chang.[14] Helicase activity was measured in 20 µl of reaction mixture containing 0.13 pmol of purified protein (=20 ng of full length protein), 25 fmol [^32^P]-labeled RNA duplex,[14] 17 mM HEPES-KOH pH 7.5, 150 mM NaCl, 1 mM MgCl2, 2 mM DTT, 1 mM spermidine, 0.3% PEG8000, 5% glycerol, 150 mM KCl, 20 units of RNasin™ Plus (Promega, USA), 1 mM ATP. The mixture was incubated for 1h at 37°C, mixed with 2X non-denaturing loading dye and subjected to gel electrophoresis through non-denaturing 8% PAGE (19:1) in 0.5X TBE at 4°C. Reaction products were visualized by autoradiography. For more information see Supplementary data.

### Generation of a HEK293T DHX30 stable knockout line

HEK293T DHX30 deficient cells were generated by transfecting a plasmid (pLentiCRISPR v2, GenScript, #52961) encoding a single guide RNA (CGAGTGCTAGCTGATCGCTT) targeting exon 7, the Cas9 endonuclease and a puromycin resistance gene under the control of the EFS promoter. Cells were transfected with TurboFect transfection reagent (Thermo Scientific) and cells were selected by puromycin for 3 days. Cells were then subjected to single cell sorting using BD FACSAria™ IIIu Cell Sorter (BD Biosciences). Single-cell clones were grown in 96-well plates for two weeks and then expanded into 6 well dishes. DHX30 knockout efficiency was assessed by Western Blotting.

### Stress treatment

HEK293T WT and DHX30 deficient cells were grown on glass coverslips coated with poly-L-Lysine using Dulbecco’s Modified Eagle Medium (DMEM) supplemented with 10% FBS. After 24 hours, cells were heat stressed at 43.5°C for 1 hour, fixed in 4% paraformaldehyde and permeabilized with 0.1% Triton X-100 (Sigma). Blocking was performed using 10% horse serum (HS). Rat monoclonal anti-ATXN2 (#8G3, kindly provided by Dr. S. Kindler, Human Genetics, UKE; Hamburg) was used as a primary antibody (1:10 in 2 % HS in PBS), followed by sgoat anti-rat IgG coupled to Alexa Fluor 647 (Thermo Fisher Scientific). Coverslips were mounted on glass microscope slides with ProLong Diamond Antifade Mountant with DAPI (Thermo Fisher Scientific). Immunofluorescence images were acquired using a confocal microscope (Leica TCS SP5, 63x/1.25 objective) and processed with ImageJ software.

### Construction of Tol2 plasmids

DHX30 cDNA plasmids were assembled using the Tol2 MultiSite Gateway® kit (Invitrogen, USA). Briefly, the cDNA of the wild-type DHX30 and DHX30 containing respective missense variants were amplified from the pEGFP-C3-DHX30 plasmids, using primers containing the appropriate att site sequences for BP recombination reactions. PCR products were purified and cloned into a pDONR221 donor vector using BP Clonase II enzyme mix following the manufacturer’s manual. The resulting middle entry clones pME-DHX30 were purified and verified by direct sequencing. To assemble the final expression plasmids, p5E-tuba1a promoter and pME-DHX30 were cloned into a Tol2-based destination vector, pDestTol2CG2 containing cmlc2:egfp transgenesis marker, using LR Clonase II Plus enzyme mix following the manufacturer’s instructions. The resultant pTol2pA2-cmlc2:EGFP;tuba1a:DHX30 vectors were purified and verified by direct sequencing.

### Zebrafish maintenance and manipulation

Tol2 transposase mRNA were synthesized using mMESSAGE mMACHINE™ T7 Transcription Kit (Ambion) per manufacturer’s instructions. 25ng/ul Tol2 mRNA and 25ng/ul of pTol2pA2-cmlc2:EGFP;tuba1a:DHX30 DNA were injected into 1-cell stage zebrafish embryos (Danio rerioAB strain). To investigate a potential dominant-negative effect 25ng/ul Tol2 mRNA and 25ng/ul equal mixture of pTol2pA2-cmlc2:EGFP;tuba1a:DHX30 with the respective variant DNA were injected into 1-cell stage zebrafish embryos. The embryos were raised and scored for abnormal development 1-7 days post fertilization. Zebrafish were maintained in the Zebrafish Research Facility at the University of Alabama at Birmingham using standard protocols. All fish were maintained at 28°C and kept at 14-hour light and 10-hour dark cycle under standard laboratory conditions. Zebrafish studies were performed according to the guidelines approved by the Institutional Animal Care and Use Committee of the University of Alabama, Birmingham.

### Generation of zebrafish *dhx30* stable knockout line

The zebrafish *dhx30* stable knockout line was generated using CRISPR/Cas9 with sgRNA target sequence 5’-TCAAGTTCAGCTGCACGGAT-3’ made by Integrated DNA Technologies (IDT) according to manufacturer’s protocol. The mutant contains an 8 bp deletion that shifts the translational reading frame after amino acid 90 and results in a premature stop codon at amino acid 107, compared to 1,173 amino acids for the wild-type (WT) protein. Mutant animals were genotyped and sequenced using primers 5’-ATCTTCACGCCAAAAACCTG-3’ and 5’-GACCACGGTTCAGCTCTCTC-3’. The dhx30 heterozygous mutants were outcrossed to the parental AB strain for at least two generations before use in experiments to eliminate potential off-target mutations. After each assay described below, test animals were individually genotyped using PCR with primers 5’-ATCTTCACGCCAAAAACCTG-3’ and 5’-GACCACGGTTCAGCTCTCTC-3’ and high-resolution melting (HRM) analysis as previously described.[15]

### Stress treatment and zebrafish whole-mount immunostaining

The dhx30 +/- animals were in-crossed to generate dhx30 +/+, +/- and -/- sibling progeny for heat shock and immunostaining analyses. 24-hour post fertilization embryos were dechorionated and incubated at 28°C or 42°C for 1 hour. After treatment, embryos were fixed overnight in cold 4% paraformaldehyde (PFA). Embryos were then dehydrated with acetone at −20°C for 7 minutes, washed in PBST [PBS+0.1% Tween 20], and blocked with 10% goat serum for at least 1 hour at room temperature. Thereafter, embryos were incubated with rabbit anti-TIAL-1 (Novus Biologicals, NBP1-79932; 1:200) overnight at 4°C, washed with PBST, and incubated with secondary antibody Alexa Fluor 488-conjugated goat anti-rabbit IgG (Invitrogen, A11034; 1:200) for 2 hours at room temperature. Embryos were washed with PBST, incubated with 100uM DAPI (1:500) to counterstain nuclei for 10 minutes and stored in PBS at 4°C. For imaging, stained embryos were mounted in 1% low melting agarose and imaged using a Nikon A1 inverted confocal microscope at approximately 50-μm Z-stacks at 5.6 μm intervals. The number of TIAL-1-labeled stress granules per 50 nuclei was quantified using Nikon NIS Element. After imaging, test animals were individually genotyped by PCR and HRM analysis to delineate the *dhx30* genotype.

### Behavioral assays

For each behavioral experiment, *dhx30* +/- animals were in-crossed to generate *dhx30* +/+, +/- and -/- sibling progeny.

#### 24-hour Sleep-Wake Activity

For each sleep-wake study, zebrafish larvae at 5 day-post fertilization (dpf) were chosen randomly and placed individually into each well of a flat-bottom 24-well plate. The activity of each larva was tracked for 24 hours consisting of 14-hour light and 10-hour dark using the DanioVision system (Noldus Information Technology). The average swimming distance was measured for 24 hours per 1-hour time-bins using EthoVision XT software (Noldus).

#### Social Preference Assay (SPA)

We adopted and modified a previously described social preference assay (SPA).[16] Briefly, SPA was performed using a flat-bottom 12-well plate and custom-built removable opaque dividers. The individual ‘‘test’’ animals, whose behaviors were analyzed, were placed in each of the 4 middle wells of the plate, and a WT conspecific of similar age and size was placed in a well either above or below each middle well. The activity of each test larva was tracked using the DanioVision (Noldus Information Technology) system and data analyzed using EthoVision XT software (Noldus). Before data acquisition, animals were given 5-minute habituation period. The ‘baseline’ activity of the test fish was then recorded while the opaque dividers were inserted between each well to prevent the animals from seeing each other. The dividers were then removed, allowing each test animal to view one well containing a conspecific animal and one empty well. The fish were given another 5-minute habituation period, followed by a 10-minute ‘post-baseline’ recording. For data analyses, wells containing test fish were divided into two 0.5 cm x 2.2 cm zones, one closest to the well containing a conspecific animal and one closest to the empty well. The amount of time spent by a test fish in each zone during the baseline and post-baseline periods was analyzed. The social preference of each test fish was quantified by calculating the social preference index (SPI) = (time spent in zone near the conspecific fish – time spent in zone near the empty well)/time spent in both zones as previously described. [16]

### Statistical Analyses

All cell line data (U2OS and HEK293T) are presented as mean ± SEM and analyzed by One-Way ANOVA followed by Dunnett’s multiple comparisons test or unpaired Student’s t-test as indicated in figure descriptions. All zebrafish-related data are presented as mean ± SEM and analyzed by unpaired Student’s t-test. The percentage of developmental defects observed upon overexpression of dhx30 was analyzed by the χ2 test.

## RESULTS

### Identification of likely causative variants in *DHX30*

We identified 25 individuals carrying likely causative variants in *DHX30* (Fig. 1). Of these, 12 individuals carry a previously reported heterozygous missense variant localizing within highly conserved helicase core motifs (HCMs): p.(Arg493His), p.(His562Arg), p.(Arg782Trp) (5 individuals including two half-sisters indicative of gonadal mosaicism), p.(Arg785Cys) (4 individuals) and p.(Arg785His). Further, 7 individuals have a novel heterozygous missense variant classified as either “likely pathogenic” or “pathogenic” according to The American College of Medical Genetics and Genomics (ACMG) guidelines (Table S1).[17] Indeed, each of these variants alters a highly conserved amino acid within a HCM predicted to be responsible for ATP binding and/or hydrolysis (Fig. 1 and Fig. S1). p.(Gly462Glu) identified in a single individual affects motif I, also referred to as Walker A motif, that binds γ phosphate and coordinates, together with motifs II and VI, ATP binding and hydrolysis in other DExH family members.[18, 19] p.(Ala734Asp) identified in two unrelated individuals, one of which (individual 6) appears to have mosaicism for the variant (Fig. S2), and p.(Thr739Ala) identified in a single individual, both affect motif V which regulates both ATP binding and/or hydrolysis and RNA binding.[2, 19] Three individuals carry p.(Arg782Gln), located within motif VI affecting the identical arginine residue (Arg782), that we previously reported p.(Arg782Trp),[8] which was identified here in five additional individuals.

**Figure 1.**
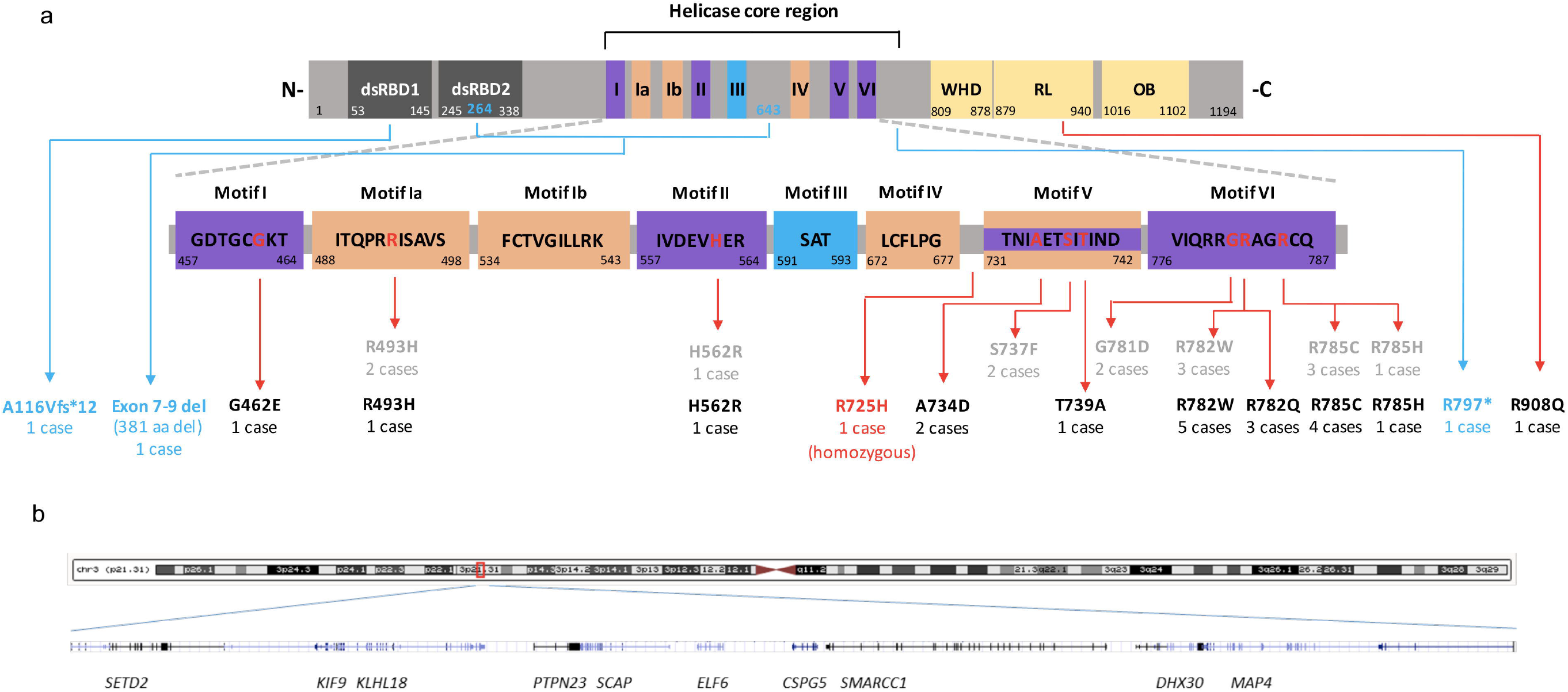
Location of identified *DHX30* germline variants. (**a**) Highly conserved sequence motifs within the helicase core region are shown with color coding that corresponds to the primary function of the motif (as previously described by Lessel *et al*., 2017). Double stranded RNA-binding domains (dsRBD) 1 and 2 at the N-terminus (N-) are shown in grey. A winged helix domain (WHD), a ratchet-like (RL) domain and an oligosaccharide binding (OB) domain are shown in yellow at the C-terminus (-C). The position of the first and last amino acid within each motif/domain is indicated below. Previously reported heterozygous missense variants and newly identified *DHX30* variants are denoted in grey and black, respectively. Frameshift and nonsense variants are denoted in blue. Mutated amino acid residues within the helicase core region are marked in red. The position of previously and newly identified mutations are indicated with red arrows. (**b**) Genomic region, chr3.hg19:g.(47098509-48109065)del, of the ∼1 Mb deletion identified in case 24.

Moreover, a homozygous variant p.(Arg725His) located within the helicase core region albeit between motifs IV and V, unlike all the missense variants mentioned above, was identified in individual 4 and classified as “variant of uncertain significance”. Additionally, a heterozygous *de novo* variant p.(Arg908Gln) was identified in individual 21. This was the only variant not located within the helicase core region; and was classified as “likely pathogenic”. Predictions based on homology to other SF2 helicases,[18] [20] and published structures of the Prp43[21] and Mle[22] revealed three novel highly conserved C-terminal regulatory domains (CTD). These include a winged helix (WH), a ratchet-like (RL) and an oligosaccharide binding (OB) fold domain (Fig. 1) with a potential role in coupling ATP hydrolysis to RNA unwinding.[23] Notably, the p.(Arg908Gln) affects a highly conserved residue within the RL domain (Fig. S1).

Furthermore we identified four individuals bearing likely pathogenic loss-of-function variants. A heterozygous *de novo* frameshift variant, p.(Ala116Val*fs*\*12) in individual 22, a heterozygous nonsense variant, p.(Arg797*) in individual 23 inherited from a mosaic mother, and a *de novo* in-frame deletion encompassing exons 7-9 of *DHX30*, leading to deletion of 381 amino acids, in individual 25. Individual 24 has a large heterozygous *de novo* deletion (arr[GRCh37] 3p21.31 (47098509_48109065)del)) encompassing ten genes including two disease genes previously associated with an autosomal dominant inheritance, *SETD2[24-26]* and *DHX30* (Fig. 1b and Fig. S3), possibly pointing to a dual diagnosis. The whole gene deletion results in haploinsufficiency, whereas the in-frame deletion, frameshift and nonsense variant, if they were to result in stable proteins, are predicted to lead to loss of functionally important domains (Fig. 1a).

Notably, none of these *DHX30* alterations were present in the gnomAD dataset v2.1.1 (Table S1),[27] indicating that they are extremely rare in the population and unlikely to be variants unrelated to disease. As previously noted *DHX30* is one of the most missense-intolerant genes in the human genome.[8] Furthermore, according to the gnomAD v2.1.1 dataset *DHX30* is, with a probability of being loss-of-function intolerant (pLI) score of 1 and a loss-of-function observed/expected upper bound fraction (LOEUF) score of 0.04, extremely loss-of-function intolerant.[27] Additionally, the degree of intolerance to deleterious variants of *DHX30* according to the Residual Variation Intolerance (RVI) score, which quantifies gene intolerance to functional mutations, is −1.51 (3.54^th^ percentile) and thus even lower than the average RVI score for genes involved in developmental disorders (0.56; 19.54^th^th percentile).[28, 29]

### Clinical spectrum of the *DHX30*-associated neurodevelopmental disorders

All 19 individuals harboring a heterozygous missense variant within a highly conserved motif in the helicase core domain have global developmental delay (GDD), intellectual disability (ID), severe speech impairment and gait abnormalities, similar to our initial findings.[8] In more detail, all individuals had intellectual disability, only nine (47%) learned to walk, all with an ataxic gait. The majority had no speech (74%), four individuals spoke only single words, and only individual 6, who is mosaic for the *de novo* p.(Ala734Asp) variant, spoke simple sentences. It is worth noting that the individuals highly benefit from communication devices (tablets, smartphones and eye-driven tablet communication systems) which significantly reduced frustration related behavior (D.L. personal communication with legal guardians and family members). Additional phenotypic features included muscular hypotonia in eighteen (95%), feeding difficulties in sixteen (84%), microcephaly in thirteen (81%, 13/16), joint hypermobility in fourteen (74%), structural brain anomalies in eleven (65%, 11/17), sleep disturbances in nine (47%), strabismus in eight (42%), autistic features in five (33%, 5/15) and seizures in four (21%) individuals (Table 1, Table S1 and supplementary data). Noteworthy, individual 6 had a relatively milder clinical course, with a moderate intellectual disability, independent walking at 2 years and 8 months and the ability to speak in simple sentences at the age of 15 years. This individual’s presentation is similar to the four individuals (#22, #23, #24 and #25), who carry either a frameshift or nonsense variant, whole-gene deletion or in-frame deletion, respectively, who all learned to walk in the second year of life, had a mild muscular hypotonia and spoke at least 20 words by the age of 3 years. Although some individuals displayed some dysmorphic features (supplementary data) we did not observe a recognizable facial gestalt, similar to our previous findings.[8]

**Table 1:** Clinical features in 25 individuals bearing pathogenic *DHX30* variants and frequency of these features in previously reported individuals.

| ***DHX30* variant** | **Heterozygous missense variants within a HCM (this study)** | **p.(Ala734Asp) mosaic (this study)** | **Haploinsufficiency / protein truncating variants (this study)** | **Homozygous p.(Arg725His) (this study)** | **Heterozygous p.(Arg908Gln) (this study)** | **Heterozygous missense variants within a HCM (previous studies: Lessel *et al.* 2017 & Cross *et al.* 2020)** |
| --- | --- | --- | --- | --- | --- | --- |
| Sex | 11 female / 7 male | female | 1 female / 3 male | male | female | 8 female / 6 male |
| Intellectual disability | 18 / 18 | + | 4 / 4 | ? | - | 13 / 13 |
| Speech ability | 14 / 18 non-verbal<br>4 / 18 single words | simple sentences | 20 word to normal speech ability | ? | normal speech ability | 11 / 13 non-verbal<br>2 / 13 single words |
| Motor development delay | 18 / 18 | + | 4 / 4 mild | + | - | 14 / 14 |
| Muscular hypotonia | 17 / 18 | + | 3 / 4 | + | - | 14 / 14 |
| Gait abnormalities | 10 / 18 no independent walking<br>8 / 18 ataxic | ataxic gait | 0 / 4 no independent walking<br>3 / 4 ataxic gait | ? | ataxic gait | 7 / 13 no independent walking<br>6 / 13 ataxic gait |
| Feeding difficulties | 15 / 18 | + | 1 / 4 | + | - | 11 / 14 |
| Microcephaly | 13 / 15 | + | 0 / 4 | - | - | 7 / 10 |
| Joint hypermobility | 13 / 18 | + | 1 / 3 | - | - | 6 / 14 |
| Brain MRI anomalies | 11 / 17 | - | 2 / 3 | + | + | 10 / 14 |
| Sleep disturbance | 8 / 18 | + | 2 / 3 | + | - | 7 / 12 |
| Strabismus | 8 / 18 | - | 2 / 4 | - | + | 6 / 14 |
| Autistic features | 4 / 14 | + | 0 / 3 | ? | - | 7 / 12 |
| Seizures | 3 / 18 | + | 2 / 3 | severe | - | 3 / 14 |

+, present; -, absent; ?, too young to evaluate; NA, unknown

Two individuals (#4 and #21) clearly stand out phenotypically. Individual 4 was homozygous for p.(Arg725His), developed early-onset infantile epileptic encephalopathy and died at 11 months. In contrast, individual 21, who harbors the *de novo* p.(Arg908Gln) variant, had unremarkable psychomotor development until the age of 8 years when she presented with progressive balance impairment with truncal ataxia. Subsequently, she experienced a decline in motor skills and developed cognitive problems with reduced concentration (Table 1, Table S1 and supplementary data).

### Effect of novel *DHX30* missense variants on ATPase activity

To corroborate the pathogenicity of the novel missense variants identified in this study, along with the recently reported p.(Ser737Phe),[11] we performed several previously established functional assays.[8] First we analyzed the ATPase activity of wild-type (WT) and mutant forms of DHX30. As previously shown, DHX30-WT acts as an RNA-dependent ATPase, and its ATPase activity is strongly stimulated by addition of RNA.[8] In contrast, and similar to the previously analyzed mutants[8] all missense variants (p.(Gly462Glu), p.(Arg725His), p.(Ala734Asp), p.(Ser737Phe), p.(Thr739Ala), p.(Arg782Gln) and p.(Arg908Gln)) show a significant reduction in ATPase activity in the presence of exogenous RNA (Fig. 2a). For control experiments, we included two common non-synonymous *DHX30* variants found in public repositories. Namely, p.(Val556Ile) located within the helicase core region albeit not within a HCM, similar to the p.(Arg725His), and p.(Glu948Lys) in the vicinity of p.(Arg908Gln). Notably, in comparison to the missense variants identified in affected individuals the ATPase activity was not significantly reduced neither for p.(Val556Ile) nor for p.(Glu948Lys) (Fig. 2b).

**Figure 2.**
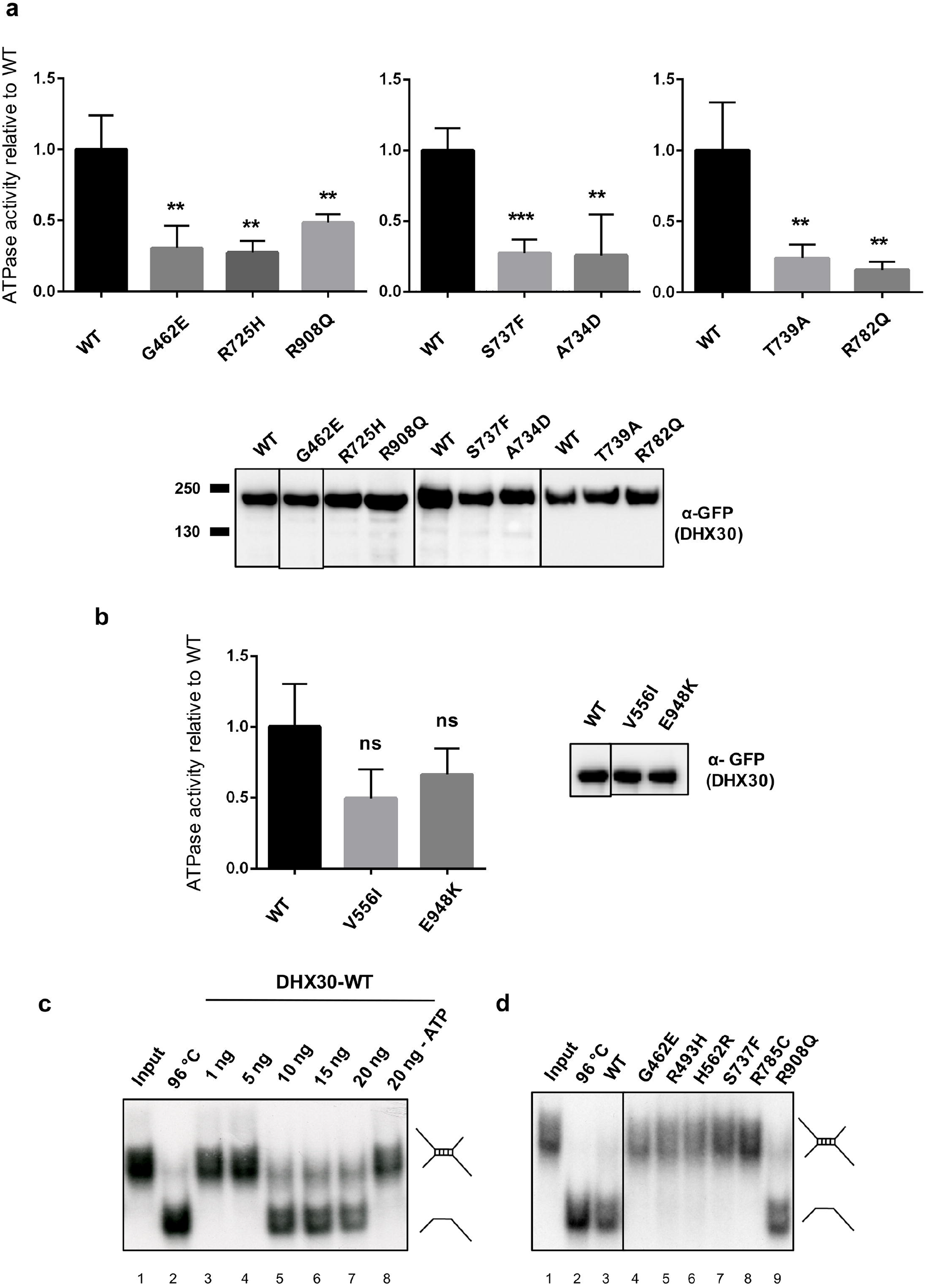
Protein variants of *DHX30* affect ATPase and helicase activity. (**a and b**) ATPase assays were performed for DHX30-WT, novel DHX30 missense variants (**a**) and two common polypmorphisms, p.(Val556Ile) and p.(Glu948Lys) (**b**) in the presence of exogenous RNA. ATPase activity was calculated by subtracting phosphate values obtained with GFP alone from those obtained with GFP-tagged DHX30-WT and mutants. These figures were then normalized on precipitated protein amounts using the optical density values of the GFP signal in the western blot. Means ± Standard Deviation values are based on 3 replications. **,***: significantly different from DHX30-WT, ns: not significantly different from DHX30-WT (**p<0.01;***p<0.001; n=3; One-Way ANOVA, followed by Dunnett’s multiple comparisons test). Values were normalized on DHX30-WT ATPase activity obtained in the presence of RNA. (**c**) Increasing amounts of His_6_-SUMO-tagged DHX30 WT protein were incubated with a ^32^P-labelled RNA substrate in the presence (lane 3-7) or absence (lane 8) of ATP and analyzed by native PAGE. The position of the RNA duplex and the single-stranded RNA are indicated in the first and second lane respectively. Their schematic representation is shown at the right side. (**d**) Helicase assay was repeated for selected DHX30 missense variants affecting either conserved motifs within the helicase core region (lane 4-8) or the auxiliary RL domain (lane 9).

### RNA helicase activity of *DHX30* is disrupted by missense variants within the helicase core motifs

DHX30 has been classified as an RH due to the presence of the highly conserved motifs in its helicase core region and sequence similarity to other RHs. To confirm that it indeed possesses an RNA helicase activity we established an RNA unwinding assay for recombinant full length DHX30 purified from bacteria as a His_6_-SUMO-tagged protein. As a substrate we used a synthetic [^32^P]-labeled RNA molecule which carries a sequence with strong propensity to self-anneal and form a double helix. Analysis of this RNA substrate by non-denaturing PAGE resulted in a single band of low electrophoretic mobility, corresponding to the dimer linked by the double helical segment. This dimer could be resolved into a band of higher mobility, the monomer, by pre-incubation at 96°C (Fig 2c). To identify the amount of the DHX30-WT necessary to resolve the dimeric form we performed a titration analysis from 1ng to 160ng. In the presence of ATP, 10ng of DHX30-WT was sufficient to resolve the dimer into the monomeric form, confirming that DHX30 indeed possesses the ATP-dependent RNA helicase activity (Fig. 2c and Fig. S4). We next analyzed the impact of selected missense variants on the helicase activity, each affecting a different helicase core motif (p.(Gly462Glu) in motif I, p.(Arg493His) in motif Ia, p.(His562Arg) in motif II, p.(Ser737Phe) in motif V and p.(Arg785Cys) in motif VI) along with p.(Arg908Gln) located in the RL domain. All missense variants within a HCM failed to unwind the RNA substrates in this assay, whereas the p.(Arg908Gln) mutant behaved similarly to DHX30-WT (Fig. 2d). It is worth noting that we subsequently failed to purify the p.(Arg725His) mutant protein product, a finding that likely suggests misfolding of this mutant protein, either due to the deposition of the insoluble protein in inclusion bodies or its direct degradation,[30] and thus could not analyze its impact on the helicase activity.

### Subcellular localization and effect on global translation of novel *DHX30* missense variants

We have previously shown that the expression of mutant forms of DHX30 induces the formation of stress granules, concomitant with a global down-regulation of translation.[8] Therefore, we repeated this analyses for selected novel missense variants. In keeping with the previous results,[8] we observed that mutants within a HCM, p.(Gly462Glu),p.(Ala734Asp),p.(Ser737Phe) and p.(Thr739Ala), also strongly accumulated in cytoplasmic foci shown to be stress granules upon co-staining with Ataxin-2 (ATXN2). Expression of the p.(Arg908Gln) mutant, however, resulted in localization to cytoplasmic aggregates that co-stained with Ataxin-2 in only 50% of the transfected cells. In contrast, p.(Arg725His) was mostly diffusely localized throughout the cytoplasm similar to the DHX30-WT (Fig. 3 and Fig. S5). Global translation was measured by incorporation of puromycin into nascent peptide chains, which were visualized with a puromycin-specific antibody. Interestingly, expression of both the HCM mutants and the p.(Arg908Gln) mutant resulted in dramatically decreased puromycin incorporation, suggestive of global decrease in protein synthesis (Fig. 3). Analogous to the results of ATPase activity, the two common variants p.(Val556Ile) and p.(Glu948Lys) were diffusely localized throughout the cytoplasm, resembling the DHX30-WT (Fig. S5).

**Figure 3.**
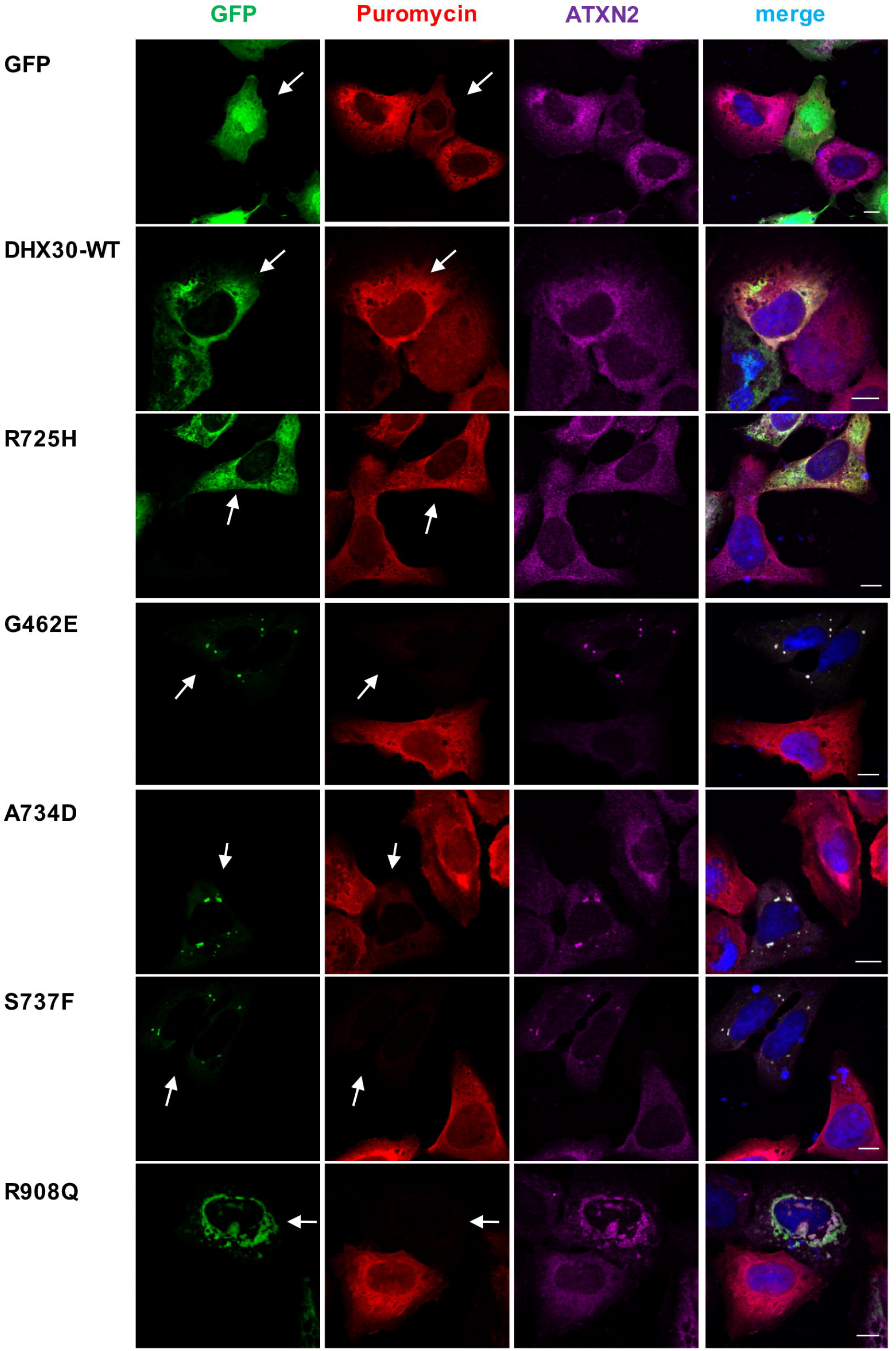
Missense variants in *DHX30* initiate the formation of cytoplasmic aggregates and impair global translation. Puromycin incorporation assay in U2OS cells expressing DHX30-GFP fusion proteins (green). Translation was monitored by staining against puromycin (red), SGs were detected by ATXN2 (magenta) and nuclei via DAPI staining (blue). Arrows indicate transfected cells. Note the correlation between formation of clusters and lack of puromycin staining. Scale bars indicate 10 µm.

### *In-vivo* analyses of selected *DHX30* missense variants

Given the somewhat conflicting results of functional analyses of the p.(Arg725His) and p.(Arg908Gln) variants, and in order to gain a better understanding of the impact of *DHX30* missense variants *in vivo*, we utilized a zebrafish model. Previous studies showed that overexpression of pathogenic alleles in zebrafish results in defective embryonic development.[31, 32] Thus, we overexpressed human wild-type DHX30 cDNA or DHX30 cDNA harboring selected missense variants, p.(Arg493His), p.(Arg725His), p.(Arg785Cys) and p.(Arg908Gln) as well as p.(Val556Ile) and p.(Glu948Lys) in zebrafish using Tol2 transposition. Tol2 mRNA and pTol2pA2-cmlc2:EGFP;tuba1a:DHX30 were co-injected into 1-cell stage zebrafish embryos. For analyses we selected embryos with strong cmlc2:EGFP expression which indicates a high level of transgene integration in somatic cells. Overexpression of DHX30-WT, p.(Val556Ile) or p.(Glu948Lys) had little or no impact on zebrafish embryonic development: over 88% of embryos displayed normal development and morphology. However, expression of DHX30 harboring one of the missense variants resulted in developmental defects in 75-90% of embryos (Fig. 4 and Fig.S6), suggesting that these mutant alleles interfere with normal embryonic development and supporting the pathogenicity of p.(Arg725His) and p.(Arg908Gln).

**Figure 4.**
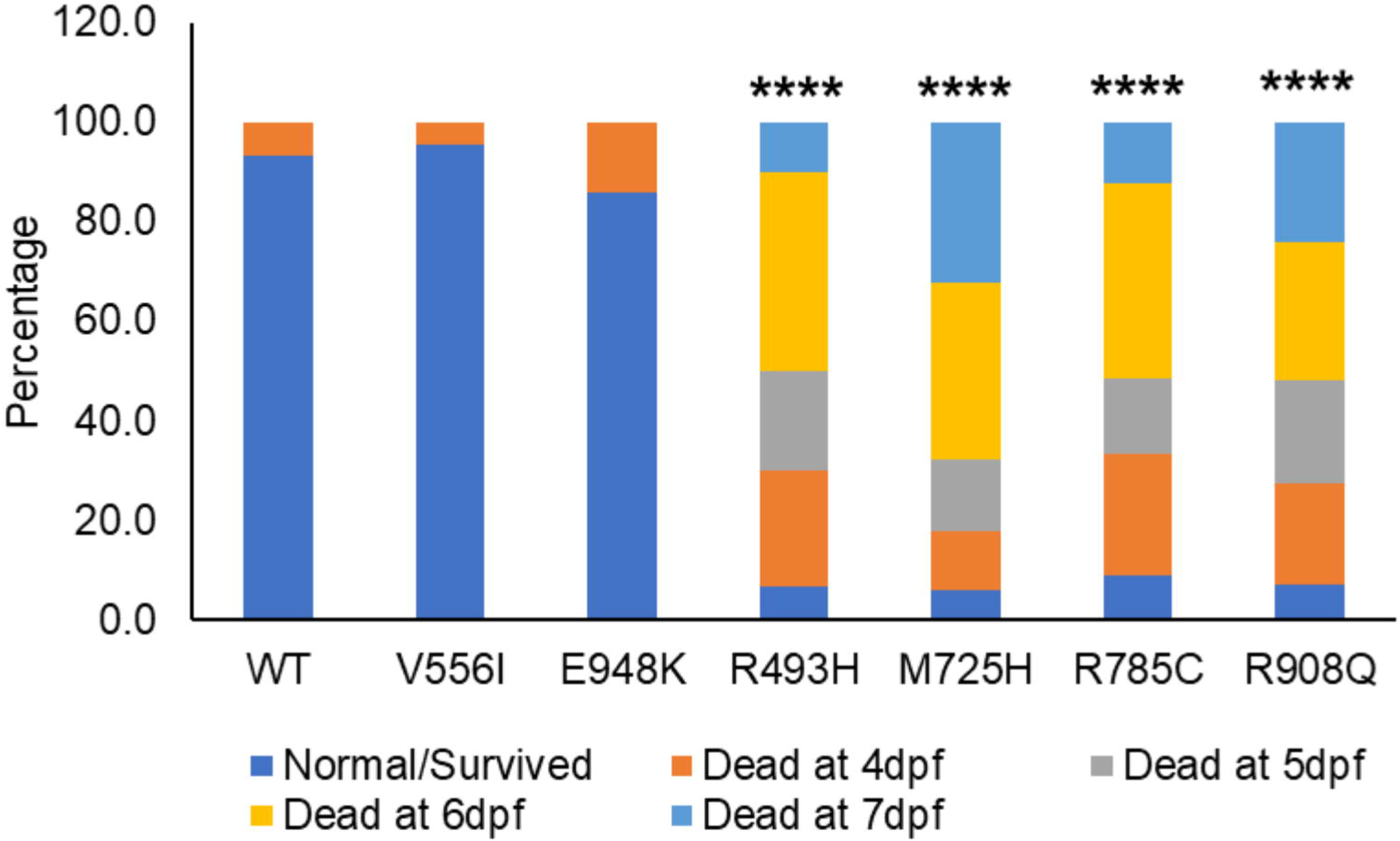
Protein variants of *DHX30* lead to embryonal developmental defects in zebrafish. In-vivo analyses of selected DHX30 missense variants. Assessment of embryonic development after injection of DHX30 WT and mutant cDNAs in a zebrafish model. Bar graph indicating the percentage of cmcl2-GFP positive zebrafish embryos assessed 4-7 days post fertilization (dpf). The presented data are derived from three independent studies. The total number of embryos assessed are 45, 23, 21, 30, 34, 33 and 29 for WT, V556I, E948K, R493H, M725H, R785C, and R908Q, respectively. ****: significantly different from WT (****p<0.0001; χ2 test)

### Analyses of the nature of the *DHX30* missense variants

Given the somewhat milder clinical presentation of individuals carrying a whole gene deletion, in-frame deletion, frameshift or nonsense variant as compared to the individuals harboringa *de novo* missense variant in one of the HCMs, we further investigated the nature of the latter. First, we analyzed the localization of the RFP-tagged DHX30-WT co-expressed with respective GFP-tagged missense variants. Notably, their equimolar expression resulted in each case in DHX30-WT being localized in the Ataxin-2 positive cytoplasmic cluster (Fig. 5). These data suggest that these missense variants either exert a dominant negative effect on the wild-type or lead to a gain-of-function since both overexpressed DHX30-WT and endogenous DHX30 are recruited to cytoplasmic clusters only after stress. [8]

**Figure 5.**
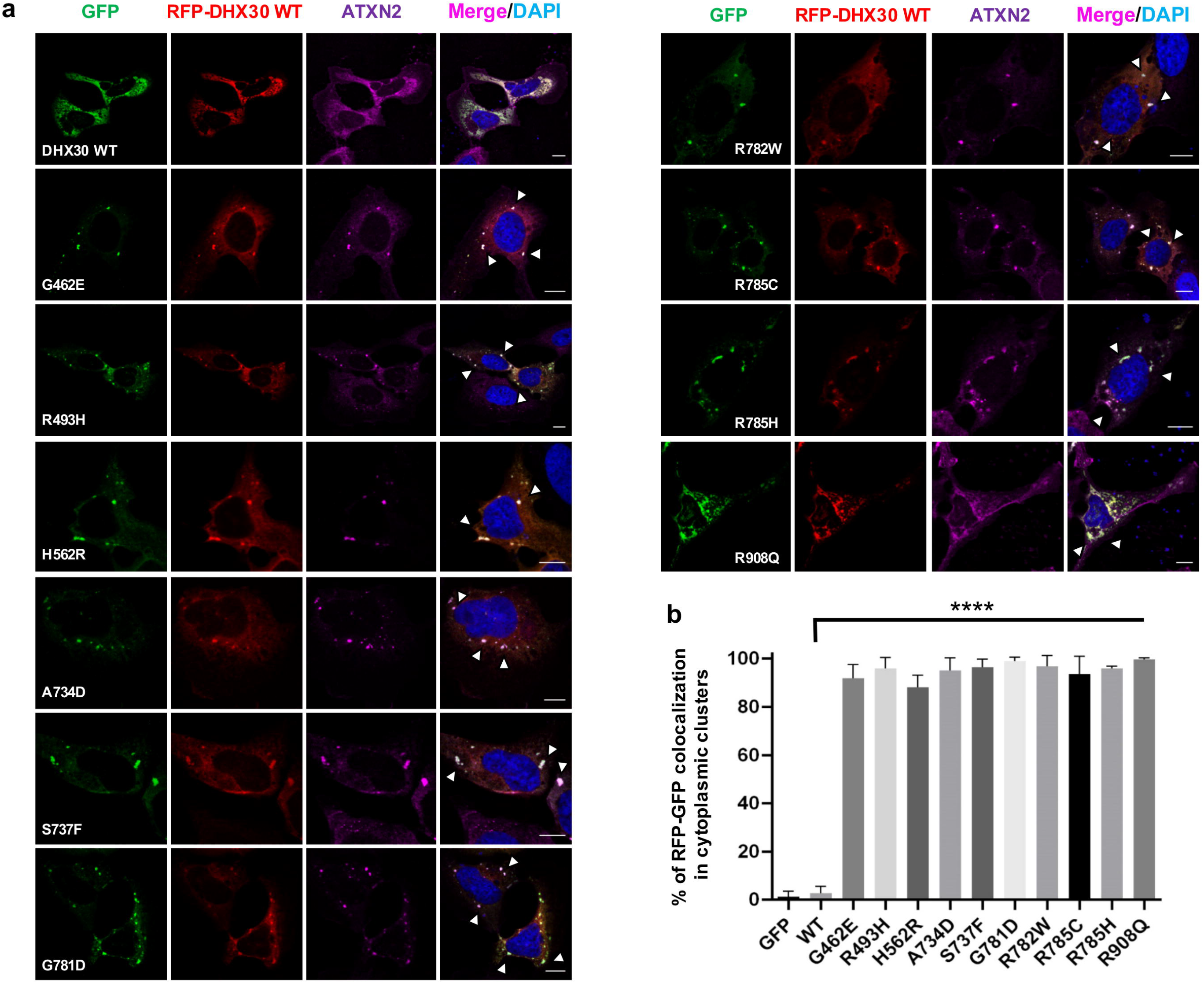
Recombinant protein variants of DHX30 induce translocation of the DHX30-wt in the cytoplasmic clusters. Immunocytochemical detection of RFP-DHX30 WT (red) and ATXN2 (magenta) after co-expression of DHX30-GFP mutants (green) in U2OS cells. Bar graph indicating the percentage of cells where RFP-DHX30 WT co-localizes with DHX30-GFP mutants within cytoplasmic clusters identified as SGs via co-staining with ATXN2 (****: significantly different form DHX30-WT: **** p< 0.0001; n > 100 from 3 independent transfections; One-Way ANOVA followed by Dunnett’s multiple comparisons test). Scale bars indicate 10 µm.

Next, we analyzed if the DHX30-WT can rescue the inability of p.(Arg493His), p.(His562Arg) and p.(Arg785Cys) to unwind the RNA. Addition of DHX30-WT to p.(His562Arg) and p.(Arg785Cys) efficiently resolved the dimer into the monomeric form even in the presence of increased amounts of the respective mutants. However, we observed only a partial rescue when DHX30-WT was added to the p.(Arg493His) variant(Fig. 6a), suggesting that the mutants cause a loss of helicase function rather than having a dominant negative effect. Given these somewhat contradictory results we turned again to the zebrafish model. We co-injected pTol2pA2-cmlc2:EGFP;tuba1a:DHX30 p.(Arg493His) or p.(Arg785Cys) with wild-type DHX30 cDNA and assessed embryonic development. Interestingly, co-injection of DHX30-WT, at similar level, partially rescued the abnormal phenotypes associated with both p.(Arg493His) and p.(Arg785Cys) (Fig. 6b), a finding that could potentially support both loss-of-function and a dominant negative effect as a mechanism underlying disease.

**Figure 6.**
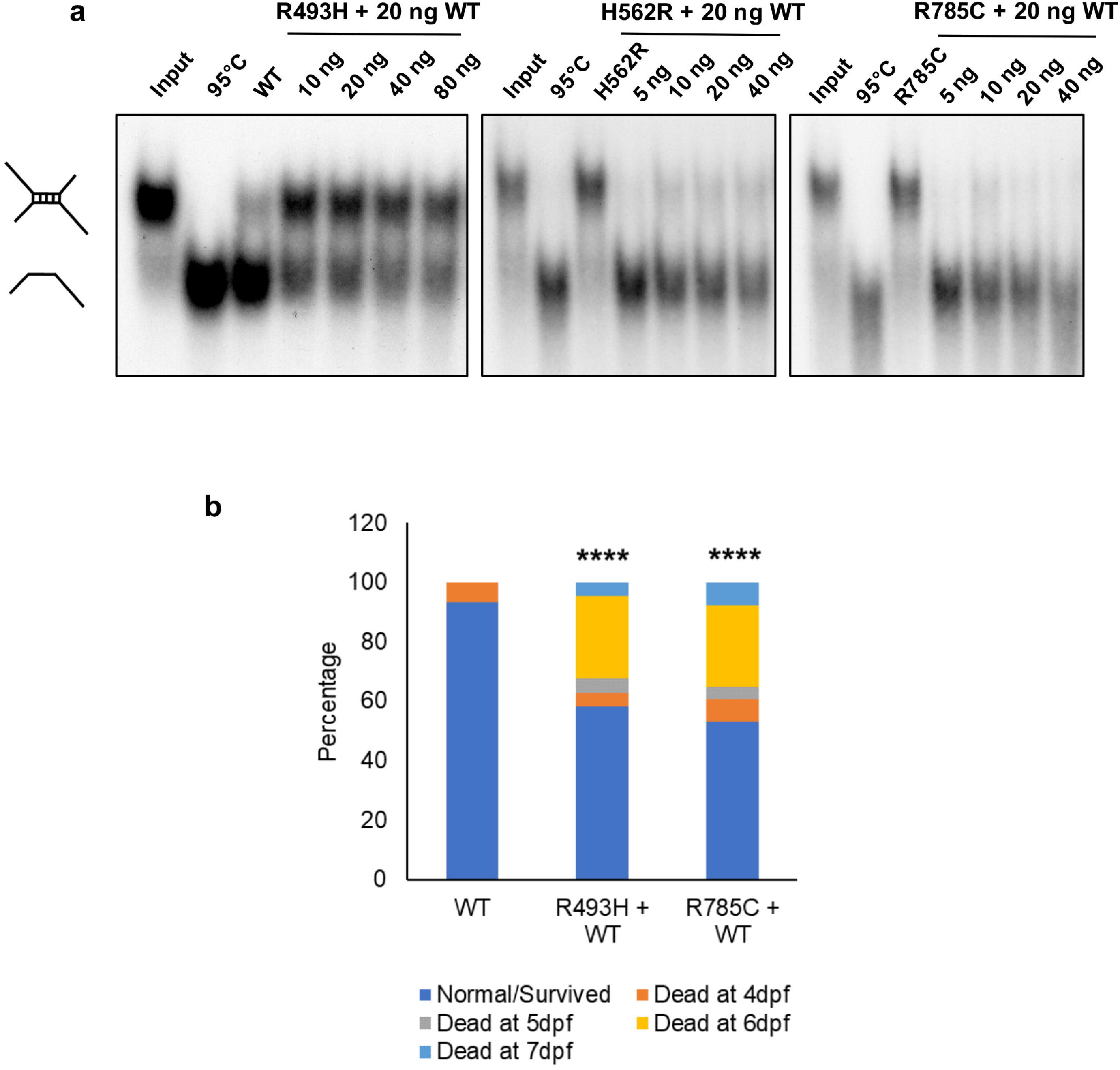
Analyses of the nature of missense variants within the helicase core motifs (HCM). (**a**) RNA unwinding activity of purified DHX30 R493H, H562R and R785C mutants was analyzed upon addition of DHX30 WT protein. Increasing amounts of mutant proteins were incubated with 20 ng of WT protein and assayed for their ability to unwind a radiolabeled RNA duplex in the presence of ATP. (**b**) Assessment of embryonic development after co-injection of DHX30 R493H and R785C with DHX30 WT cDNA in a zebrafish model. Bar graph indicating the percentage of cmcl2-GFP positive zebrafish embryos 4-7 days postfertilization (dpf). The presented data are derived from three independent studies. The total number of embryos assessed are 58, 43, and 51 for WT, R493H+WT, and R785C+WT, respectively. ****: significantly different from WT (****p<0.0001; χ2 test)

### DHX30 deficiency impairs stress granules formation in HEK293T cells and zebrafish

To further characterize the role of DHX30 we established HEK293T DHX30 stable knockout lines. CRISPR/Cas9 based knockout (KO) of DHX30 in HEK293T cells yielded several cell lines with a residual DHX30 immunoreactivity of less than 10 % (Fig. 7). Given that DHX30 is recruited to SGs, we wondered whether DHX30 additionally plays a role in SG formation. Therefore, we assessed the ability of KO cells to induce SGs or cytoplasmic clusters following heat stress treatment. By incubating cells at 43.5°C, a condition after which endogenous DHX30 accumulates in SGs,[8] we observed that KO cells had a significantly reduced number of SG-positive cells as compared to HEK293T WT cells (Fig. 7). These data suggest a previously unknown role of DHX30 in SG assembly. Combined with our previous findings (Fig. 3 and 5) these data actually suggest that the HCM missense variants exhibit a gain of function by triggering SG formation which results in global translation inhibition.

**Figure 7.**
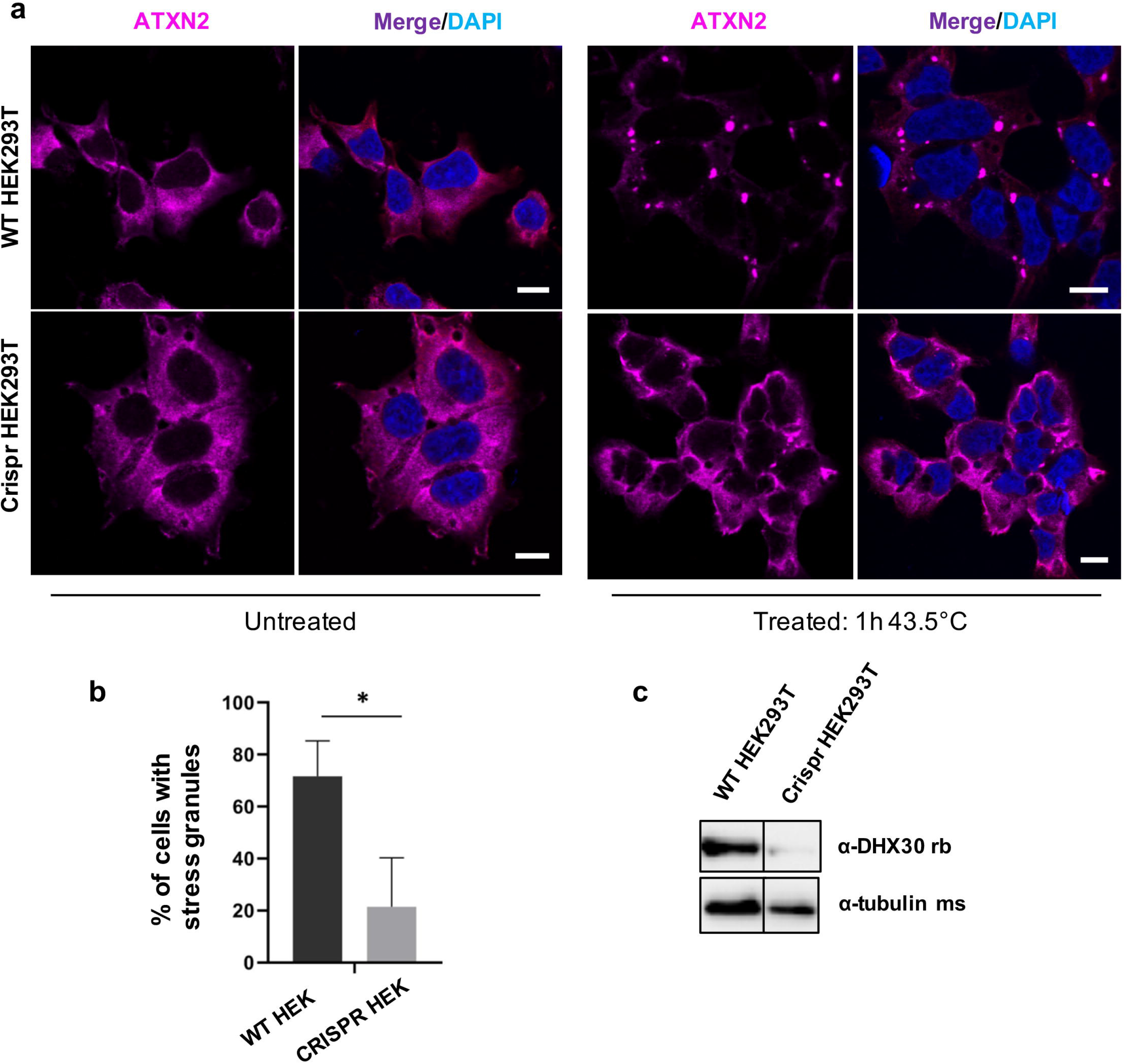
DHX30 deficiency in HEK293T cells leads to reduced formation of stress granules. (**a**) Immunocytochemical detection of endogenous ATXN2 (magenta) in WT HEK293T cells and DHX30 deficient HEK293T cells before (left panel) and after (right panel) heat shock at 43.5°C for 1h. Note that, upon heat stress and depletion of DHX30 (right hand, lower panel), ATXN2 does not alter its diffuse cytoplasmic distribution to accumulate in cytoplasmic foci, as observed in WT HEK293T cells (right hand, upper panel). Nuclei are identified via DAPI staining (blue). Scale bars indicate 10 µm. (**b**) Bar graph indicating the percentage of cells containing stress granules. (*: significantly different from WT HEK293T cells: *p<0.05; n > 200 from 3 independent experiments; Unpaired t-Test). (**c**) Western blotting detection of DHX30 knock-out efficiency in HEK293T cells. Expression of DHX30 was reduced by 90% as detected by a DHX30 specific antibody. Tubulin was used as loading control.

Next, we generated a predicted null allele in the single zebrafish dhx30 ortholog using CRISPR/Cas9. At day five post fertilization, transcript levels of dhx30 were barely detectable in homozygous mutant animals compared to wild-type, whereas heterozygous siblings displayed ∼30% lower dhx30 expression as compared to wild type, potentially due to nonsense mediated decay of the mutated alleles (Fig. S7). The homozygous mutant animals are viable, fertile, and morphologically indistinguishable from their wild-type and heterozygous siblings (data not shown). Previous studies have demonstrated that during early embryonic development, zebrafish exhibit robust SG formation in response to stress, such as heat shock.[33] Therefore, based on our *in vitro* findings, we first asked whether dhx30 mutant zebrafish also exhibit impaired SG formation *in vivo*. At 24 hour-post fertilization and normal condition, compared to dhx30-WT the homozygous mutant exhibited significantly lower number of SGs, determined by staining for TIAL-1, an established stress granule marker (Fig. 8). Although an increase in SG formation occurred upon heat shock, the number of TIAL-1-labeled SGs remained significantly lower in the homozygous mutants compared to sibling controls (Fig. 8). Thus, these data show that SG formation is compromised in the homozygous mutants and suggest an evolutionarily conserved role for DHX30 in SG assembly.

**Figure 8.**
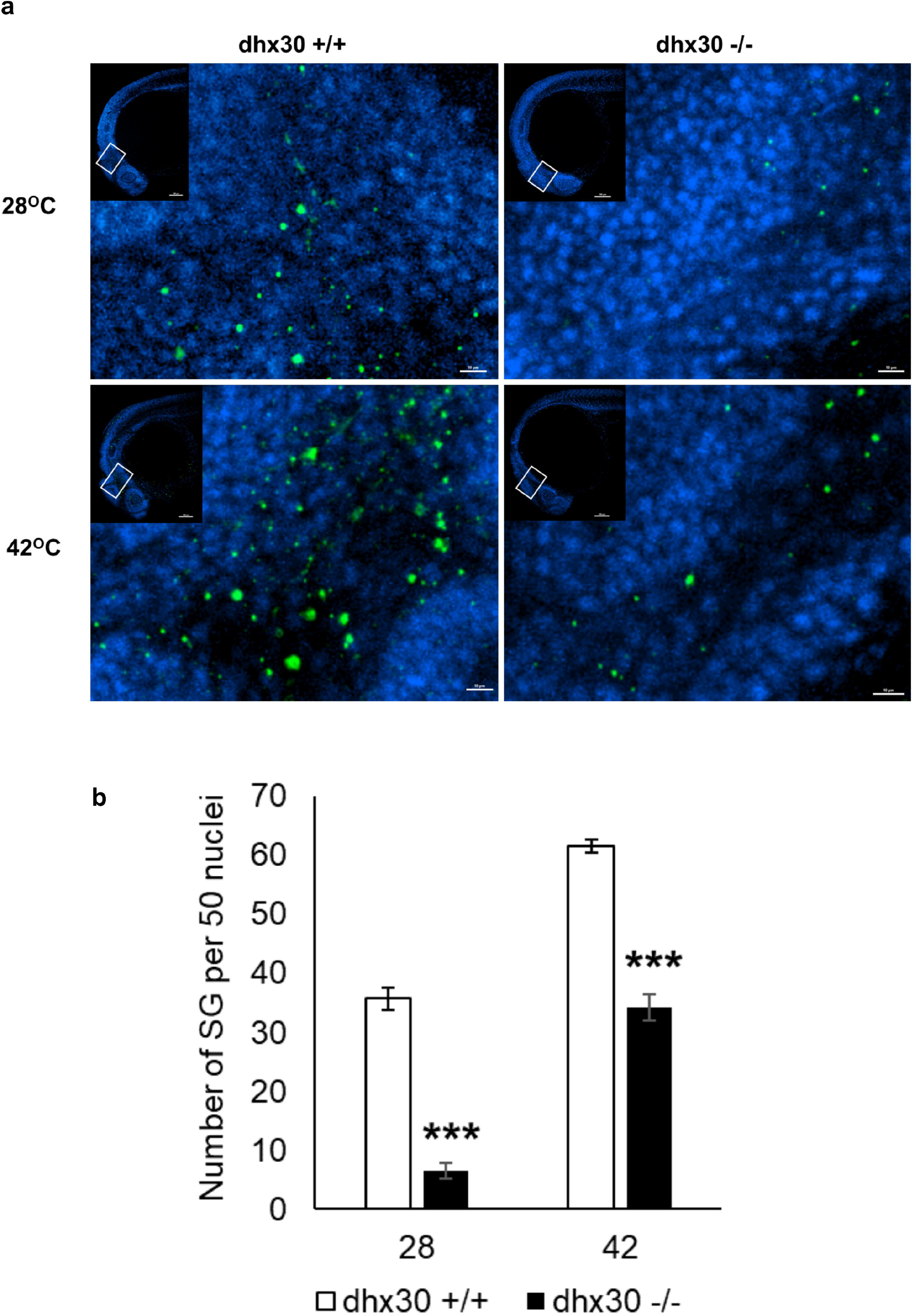
DHX30 deficiency in zebrafish cells leads to reduced formation of stress granules. (**a**) Representative confocal images of TIAL-1-labeled stress granules (green) in dhx30 wild-type (+/+) and homozygous mutants (-/-) underwent normal conditions or heat shock treatment at 42oC. Nuclei were counterstained with DAPI (blue). (**b**) Analyses of TIAL-1-labeled stress granules per 50 nuclei. The total number of embryos assessed are 8, 9, 8, and 8 for dhx30+/+ (28°C), dhx30 -/- (28°C), dhx30 +/+ (42°C), and dhx30 -/- (42°C), respectively. Data are presented as means ± standard error of mean based on the indicated number of embryos. ***: significantly different from DHX30+/+ (***p<0.001; unpaired Student’s *t*-test)

### *Dhx30*-deficient zebrafish display altered behavioral activity

We next examined whether dhx30 mutant zebrafish exhibit abnormal sleep-wake activity and social behaviors, similar to those recently observed in a zebrafish model of the NR3C2–related neurodevelopmental disorder.[16] We first analyzed sleep-wake behaviors in 5-day-old dhx30 KO mutants. Compared to wild-type and heterozygous siblings, the homozygous mutants displayed significantly less activity during the day and more nocturnal activity (Fig. 9a-c), mimicking somewhat the sleep disturbances in individuals affected by a DHX30-related neurodevelopmental disorder. Additionally, using an established social preference assay, we observed that the wild-type and heterozygous animals showed the previously described social behavior of preferring to stay close to conspecific fish of similar age and size, whereas the homozygous animals did not show this preference (Fig. 9d-e). There were no obvious dysmorphic phenotypes in the homozygous mutant animals compared to their wild-type and heterozygous siblings. We propose, therefore, that the mutant phenotype was not simply due to developmental delay but influenced by abnormalities in complex neural circuitry. Taken together, our data indicate that dhx30 KO zebrafish have a social behavioral deficit with altered sleep-wake activity, which is consistent with findings in DHX30-related neurodevelopmental disorders.

**Figure 9.**
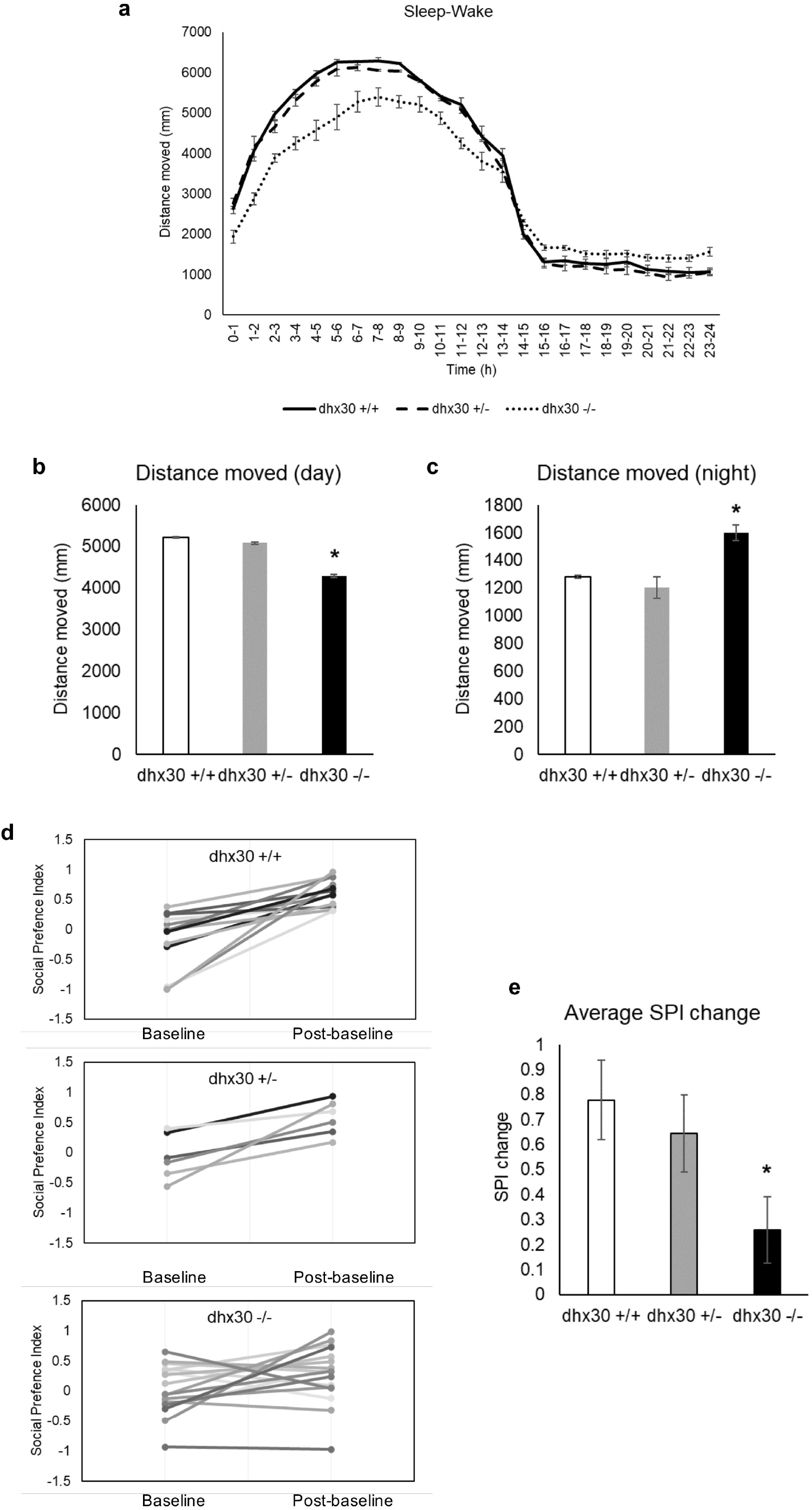
Behavioral analyses of dhx30 mutant zebrafish. (**a**) Distance moved of dhx30 mutants and wild-type sibling controls measured at 5 days post fertilization. (**b**) Average of distance moved during 14-hour daytime, and (**c**) Average of distance moved during 10-hour nighttime. N= 15, 18, and 25 for +/+, +/-, and -/- animals, respectively. (**d**) Social preference index (SPI) calculated during 10-min baseline and post-baseline period. SPI = 1 indicates a fish that spends 100% of its time near a conspecific, SPI = - 1 indicates a fish that spends 100% of its time near the empty well, and SPI = 0 indicates a fish that spends equal amounts of time near the conspecific and near the empty well. (**e**) The change in SPI between baseline and post-baseline, indicating the preference of zebrafish to stay close to conspecific fish. N= 13, 6, and 17 for +/+, +/-, and -/- animals, respectively. Data are presented as means ± standard error of mean based on the indicated number of embryos. *: significantly different from DHX30+/+ (*p<0.05; unpaired Student’s *t*-test)

## DISCUSSION

Our study has allowed further delineation of the clinical spectrum of *DHX30*-related neurodevelopmental disorders through analysis of 25 novel affected individuals, partially facilitated by the use of a social media-based family support group. Individuals harboring heterozygous missense variants affecting highly conserved residues within a HCM present with global developmental delay, intellectual disability, muscular hypotonia, severe gait abnormalities (if walking is acquired), and remain non-verbal or speak only single words. We also identified microcephaly as an additional common feature. Individuals with either a mosaic missense variant within a HCM, or with variants resulting in haploinsufficiency or with protein-truncating variants all learned to walk in the second year of life, had a mild muscular hypotonia and spoke at least 20 words by the age of 3 years. Therefore, based on the clinical and molecular findings we suggest a classification in two *DHX30*-associated neurodevelopmental disorder subtypes.

It is worth noting that the identified heterozygous deletion in individual 24 also encompasses the first 15 *SETD2* exons, suggestive of a dual diagnosis. However, given the phenotypic differences in the seven individuals reported to date, some of whom inherited their *SETD2* variant from an apparently unaffected parent,[24-26], we are unsure to what extent loss of *SETD2* contributed to the phenotype observed in this individual.

Identification of affected individuals with a milder phenotype challenges naming of this disorder “Neurodevelopmental disorder with severe motor impairment and absent language” (NEDMIAL; OMIM # 617804). Notably, only 9 of the 25 individuals (36%) presented here had a severe motor impairment (never learned to walk) and 9 out of 25 individuals (36%) spoke at least single words, thus did not have a completely absent language. Therefore we suggest referring to these conditions as *DHX30*-associated neurodevelopmental disorders in the future.

To provide further evidence for the pathogenicity of the novel *DHX30* variants and gain better insight into the genotype-phenotype correlation we performed several *in vitro* and *in vivo* analyses. For this, we have now formally confirmed that DHX30 possesses ATP-dependent RNA helicase activity. In line with their absence from public databases and high evolutionary conservation of affected amino acid residues, all novel missense variants within a HCM resulted in impaired ATPase activity (all were within an ATP binding and hydrolysis motif), impaired helicase activity, and showed an increased propensity to trigger stress granule (SG) formation resulting in inhibition of global translation, as expected from the previous study.[8] In addition, selected HCM missense variants interfere with normal zebrafish embryonic development.

We have previously suggested that the missense variants within HCM might have a more severe effect than a loss of one gene copy.[8] This hypothesis is now supported by identification of four affected individuals carrying variants that result in either haploinsufficiency or a truncated protein, all of whom presented with a milder phenotype as compared to the individuals harboring missense variants within HCM. To gain further insight into the nature of these variants we determined that DHX30-wt can rescue the inability of selected HCM missense variants to unwind an RNA duplex, and that co-injection of DHX30-wt with selected HCM missense variants can partially ameliorate the observed zebrafish phenotypes. These data point to loss-of-function effects of the HCM mutants on a molecular level. However, co-expression of HCM missense variants together with the DHX30-WT, resulted in recruitment of DHX30-WT into SG’s, a finding that might possibly suggest a dominant negative effect. However, it is worth noting that DHX30-WT, as well as the endogenous protein, are recruited to the SG’s after stress induction.[8] Thus, HCM missense variants might actually result in a detrimental gain-of-function by inducing SG formation with concomitant global translation impairment even without endogenous or exogenous stressors.

To gain further clarity we focused on the relation of DHX30 to SG formation. Using CRISPR/Cas9 based technology, we established two DHX30 knockout models. Analyses of both, DHX30 deficient HEK293T cells and zebrafish, revealed an impairment of SG formation upon heat stress, pointing to an essential and evolutionary conserved role of DHX30 in SG assembly. These findings provide a molecular explanation for the abovementioned phenotypic differences, as they strongly suggest that pathogenic missense HCM variants, in addition to the loss of ATPase or RNA-binding activity and with impaired helicase function, exert a selective gain-of-function by triggering SG formation. This is in line with our hypothesis that due to SG hyper-assembly these pathogenic variants generate a chronic condition of impaired translation.[8] Noteworthy, impaired translation due to aberrant SG formation is associated with a broad variety of neurodegenerative and neurodevelopmental diseases.[8, 34] Furthermore, repeat expansion underlying *C9orf72*-associated neurodegenerative disorders was recently suggested to result in chronic cellular stress due to aberrant SG formation.[35]

Beyond providing the molecular explanation for the genotype-phenotype correlation of these two subtypes we additionally performed *in vivo* behavioral modeling of zebrafish dhx30 KO’s. Zebrafish exhibit all the hallmarks of mammalian sleep by utilizing neurotransmitters known to coordinate sleep and wake states in humans.[36] Analysis of dhx30 deficient animals revealed a compromised sleep/wake behavior, as they were less active during the day but more active and slept less at night than dhx30-WT animals. This is partially reminiscent of the sleep disturbances observed in almost half of DHX30-affected individuals. Additionally, homozygous dhx30 KO animals displayed altered social behavior as manifested by their performance in the social preference assay, e.g. showing reduced preference for conspecifics as compared to dhx30-wt zebrafish. The observed social behavioral deficits and altered sleep-wake activity are similar to the findings in zebrafish models of other neurodevelopmental disorders,[16, 37] and to some extent recapitulate the clinical findings in individuals affected by the DHX30-related neurodevelopmental disorder.

Furthermore, we present here two individuals who clearly stand out both in terms of their clinical presentation and their identified *DHX30* variant. Individual 4 with an early-lethal infantile epileptic encephalopathy carries a homozygous missense variant, p.(Arg725His), and individual 21 with a *de novo* p.(Arg908Gln) variant shows late-onset progressive ataxia. Trio-WES analysis performed in both individuals identified these *DHX30* variants as the only candidates (Supplementary Data). As these variants occurred outside the HCM motifs, we included two similarly located common non-synonymous *DHX30* variants found in gnomAD, p.(Val556Ile) and p.(Glu948Lys), in our functional analysis for comparison. However, these two latter variants behaved similarly to DHX30-WT in all assays performed (Table 2).

**Table 2:** Summary of functional analyses of missense variants.

| *DHX30* variant | p.(Gly462Glu), p.(His562Arg), p.(Ala734Asp),<br>p.(Ser737Phe), p.(Thr739Ala), p.(Gly781Asp)<br>p.(Arg782Gln) p.(Arg782Trp), p.(Arg785Cys),<br>p.(Arg785His) | p.(Arg493His | p.(Arg725His) | p.(Arg908Gln) | p.(Val556Ile) | p.(Glu948Lys) |
| --- | --- | --- | --- | --- | --- | --- |
| Location in DHX30 | Helicase core motifs I, II, V or VI<br>(nucleotide-interacting motifs) | Helicase core motif Ia<br>(nucleic acid-binding) | Helicase core region,<br>between motifs IV and V | ratchet-like domain | Helicase core region,<br>between motifs Ib andII | C-terminal region |
| gnomAD v2.1.1 | not identified | not identified | not identified | not identified | 0/39/282352 | 1/49/282090 |
| ATPase activity | reduced | similar to wt* | reduced | reduced | similar to wt | similar to wt |
| RNA binding capacity | n.d. | reduced* | n.d. | n.d. | n.d. | n.d. |
| Helicase activity | reduced** | reduced | n.d.*** | similar to wt | n.d. | n.d. |
| Cellular localization | stress granules | stress granules | cytoplasmic, similar to wt | cytoplasmic aggregates | cytoplasmic, similar to wt | cytoplasmic, similar to wt |
| Puromycin incorporation | impaired | Impaired* | similar to wt | impaired | n.d. | n.d. |
| Zebrafish development | impaired** | impaired | impaired | impaired | similar to wt | similar to wt |

n.d., not determined; \*, Lessel et al. 2017; \*\*, only selected variants analyzed; \*\*\*, unable to purify the protein;

For the variant p.(Arg725His), located within the helicase core region but not within a HCM, we observed a reduced ATPase activity. However, unlike HCM missense variants it does not trigger SG hyper-assembly. When attempting to analyze its impact on the helicase activity we consistently failed to purify the p.(Arg725His) mutant protein product. We therefore suggest that this biallelic variant leads to a loss-of-function, likely due to misfolding. The fact that it was inherited from unaffected heterozygous parents indicates that its effect is somewhat milder as compared to the variants identified here resulting in haploinsufficiency or protein-truncation, and that similar heterozygous missense variants may not contribute to disease.

The *de novo* missense variant p.(Arg908Gln) affects a highly conserved residue within the RL domain. This variant impairs the ATPase but not helicase activity of DHX30, suggesting that the RL domain is required for the coupling of helicase activity to ATP hydrolysis. Whereas this variant leads to formation of aberrant cytoplasmic aggregates, which cannot be eliminated by co-expression of DHX30-WT, not all of these aggregates/foci could be confirmed to be translationally silent SGs.

The functional characterization of both variants identified differences to HCM missense variants, which may potentially explain the genotype-phenotype correlation. Additionally, both individuals presented with clinical signs and symptoms observed in other affected individuals, suggestive of a phenotypic continuum. We cannot exclude, however, the possibility that they carry additional variants with phenotypic consequences, which were undetected by trio-whole exome sequencing. Identification of similarly affected individuals carrying similar variants is required to establish their causality.

## CONCLUSION

The identification of 25 affected individuals has expanded the clinical and genetic spectrum of the *DHX30*-associated neurodevelopmental disorder. Our data suggest the existence of clinically distinct subtypes correlating with location and nature of pathogenic variants. Our study highlights the usefulness of social media-based family support groups as a resource in defining ultra-rare disorders as well as the need for *in-depth* functional characterization of potentially pathogenic variants to understand their biological consequences. We confirmed that DHX30 is an ATP-dependent RNA helicase, and showed that DHX30 is essential for stress granule assembly in cellular and *in vivo* models. Missense mutations in helicase core motifs lead to a loss of ATPase and helicase activity, concomitant with a gain-of function with respect to SG formation, and a severe phenotype. In contrast, *DHX30* loss-of-function mutations are associated with a milder phenotype. Additional studies are required to further delineate the variety of clinical outcomes underlying different *DHX30* variants as well as the roles of DHX30 in various aspects of RNA metabolism.

## Data Availability

The raw whole-exome sequencing data that support the findings in affected individual cannot be made publicly available for reasons of patient confidentiality. Qualified researchers may apply for access to these data, pending institutional review board approval.

## AUTHORS’ CONTRIBUTIONS

I.M., N.D.P.D., H.H., J.W., J.M.P., U.F., N.C.Y., H-J.K. and D.L. generated and analyzed the functional data. D.L. and H-J.K. supervised the study. D.L. wrote the manuscript. J.B.M., J.A., T.A., S.B., G.B., D.B., A.B., P.J.B., S.B., T.B., F.B., L.A.B., G.J.B., Ø.L.B, J.C., J.D., L.F.E., C.E., J.F., D.G., C.A.H., M.H., Y.H-M., G.H., A.J., L.K., B.K., C.K-B., C.Kr., C.Ku., G.L.G., U.W.L, L.M.B, J.A.M-A., M.M., D.T.M., K.Q.M., B.M., C.N., S.F.N., T.P., F.R., H.R., S.F.R., J.S-H., P.B.S, A.S., S.S., A.P.A.S., K.To., K.Tv., J.H.W., C.Z., K.M., J.J., F.Q-R., identified and collected patients. All authors have read and approved the final manuscript.

## ACKNOWLEDGEMENTS

We thank all family members for their participation and collaboration, Hans-Hinrich Hönck (Institute for Human Genetics, UKE Hamburg) for technical assistance, and UKE microscopic imaging facility (umif) for providing assistance with confocal microscopes. This work was funded in part by Werner Otto Stiftung (to D.L and H-J.K) and Deutsche Forschungsgemeinschaft (LE4223/1-1 to D.L.; Kr1321/8-2 to H-J.K), by startup funds from University of Alabama, Birmingham (to N.C.Y), by NIH U54 OD030167 (to J.P.M.), by the UCLA Pathology Translational Research Fund (to J.B.M. and F.Q-R.) and by the UCLA California Center for Rare Diseases (to S.F.N).

