## supplementary data for "Genotype–phenotype correlations and novel molecular insights into the *DHX30*-associated neurodevelopmental disorders"

**Mannucci *et al*.**

### Clinical reports:

Detailed clinical data on the affected individuals presented here are not available through the medRxiv preprint server. Qualified researchers and clinicians may apply for access to these data, pending institutional review board approval.

**Supplementary Methods**

**Expression of 6xHis-SUMO-DHX30 constructs**

6xHis-SUMO-DHX30 wild-type and mutant constructs were transformed into *E. coli* BL21 (DE3) pLysS pRARE competent cells. Colonies were inoculated into 100 mL of superbroth (SB) medium supplemented with 2% glucose, 0.1% kanamycin and 0.1% chloramphenicol and incubated overnight at 37°C under shaking at 200 rpm. The 100 ml bacterial pre-culture was transferred into 2 L of SB medium and incubated for 3h at 37°C under shaking at 180 rpm until an $OD_{600}$=1 was reached. Then, protein expression was induced by adding 1 mM of IPTG and cells were cultured overnight at 14°C under shaking at 180 rpm.
Cells were collected by centrifugation and cell pellets were resuspended in 50 mL of lysis buffer (20 mM HEPES pH 7.5, 500 mM NaCl, 5 mM $MgCl_2$, 0.01% NP-40, 10% glycerol, 5 mM β-mercaptoethanol, 0.1 mM AEBSF, 0.5 mg/L leupeptin/pepstatin A, 2 mg/L aprotinin). Solubilized samples were additionally sonified with the Brandson 250 sonifier. Insoluble cell debris was removed by ultracentrifugation with a 45Ti Beckman rotor at 25 000 rpm for 1h at 4°C.

**Protein purification.** Cell lysates were incubated for 2h at 4°C on Ni-NTA beads (Qiagen, #30210) previously equilibrated with lysis buffer. Then, beads were washed three times with 50 times bed volume of washing buffer (20 mM HEPES pH 7.5, 500 mM NaCl, 5 mM $MgCl_2$, 0.01% NP-40, 10% glycerol, 40 mM Imidazole, 5 mM β-mercaptoethanol, 0.1 mM AEBSF, 0.5 mg/L leupeptin/pepstatin A, 2 mg/L aprotinin). Bound proteins were eluted with 8 times bed volume of elution buffer (20 mM HEPES pH 7.5, 500 mM NaCl, 5 mM $MgCl_2$, 10% glycerol, 300 mM imidazole, 5 mM β-mercaptoethanol,). The elution fractions were subsequently analyzed for the presence of the target protein by SDS-PAGE and Coomassie blue staining. Afterwards the fractions of interest were pooled and sample was concentrated using concentrators (Vivaspin™ *500*, MWCO 10 000). 2 - 5 mg of the concentrated sample was further purified by size exclusion chromatography (Superdex 75) with the Äkta™ purifier system (GE Healthcare). Fractions were analyzed for the presence of the target protein by SDS-PAGE and Coomassie staining. Samples were quantified from the Coomassie stained gel using the ImageJ software.

***In vitro* synthesis of RNA molecules.** To test the RNA unwinding activity of DHX30, a $^{32}$P-Labeled RNA duplex was synthesized using the T7 RNA polymerase from a linearized DNA template designed by (Tseng-Rogenski and Chang, 2004). The *in vitro* transcription reaction mix was prepared as follows: 1X transcription buffer (40 mM Tris-HCl pH 7.9, 1 mM Spermidine, 26 mM MgCl2, 0,01% Triton X, 5mM DTT), NTPs (GTP 8 mM, ATP 5 mM, CTP 5 mM, UTP 2 mM, 50 µCi of $^{32}$P-UTP), 3 µM DNA template, 3 µM top strand primer (5'-TAATACGACTCACTATAG-3'), 7 U of T7 RNA polymerase. Transcription reactions were incubated for 2h at 37° C. RNA was precipitated by adding 0.1 volumes of 3M NaAc (pH 5.5) and 3 volumes of ethanol for 30 min at -20°C. Then, samples were centrifuged at 13 000 g, 30 min at 4°C. RNA pellets were rinsed with 70 % ethanol and resuspended in $H_2O$.
RNA samples were mixed with 2X denaturing RNA loading dye (1X Tris-Borate-EDTA pH 8.3, 95% formamide, 0.1% bromophenol blue, 0.1% xylene cyanol FF) and analyzed on 8% UREA-PAGE in 1X TBE. Radioactive signals were detected by autoradiography and the RNA band was excised from the gel.
The RNA product was extracted from the gel in RNA extraction buffer (200 mM Tris-Hcl pH 7.0, 0.1% SDS, 1 mM EDTA). Subsequently, RNA was precipitated as described above and the RNA pellets were resuspended in 100 mM KCl.
To promote the formation of the RNA duplex, the RNA sample was boiled at 95°C for 5 min and cooled down over 2h. The sample was mixed with 2X non-denaturing loading dye (1X TBE, 20% glycerol, 0.1% bromophenol blue, 0.1% xylene cyanol FF) and separated on 8% native PAGE. The RNA duplex

was excised from the gel and purified as described before. The RNA concentration was determined by measuring absorbance at 260 nm.

**RNA unwinding assay**

The helicase activity was measured in 20 µl of reaction mixture containing 0.13 pmol of purified protein (=20 ng of full length protein), 25 fmol radioactively labeled RNA duplex, 17 mM HEPES-KOH pH 7.5, 150 mM NaCl, 1 mM $MgCl_2$, 2 mM DTT, 1 mM spermidine, 0.3% PEG8000, 5% glycerol, 150 mM KCl, 20 units of RNasin™ Plus (Promega), 1 mM ATP. The mixture was incubated for 1h at 37°C and subsequently mixed with 2X non-denaturing loading dye and subjected to gel electrophoresis through non-denaturing 8% PAGE (19:1) in 0.5X TBE at 4°C. Reaction products were visualized by autoradiography.

### Supplementary Figures 1 – 7.

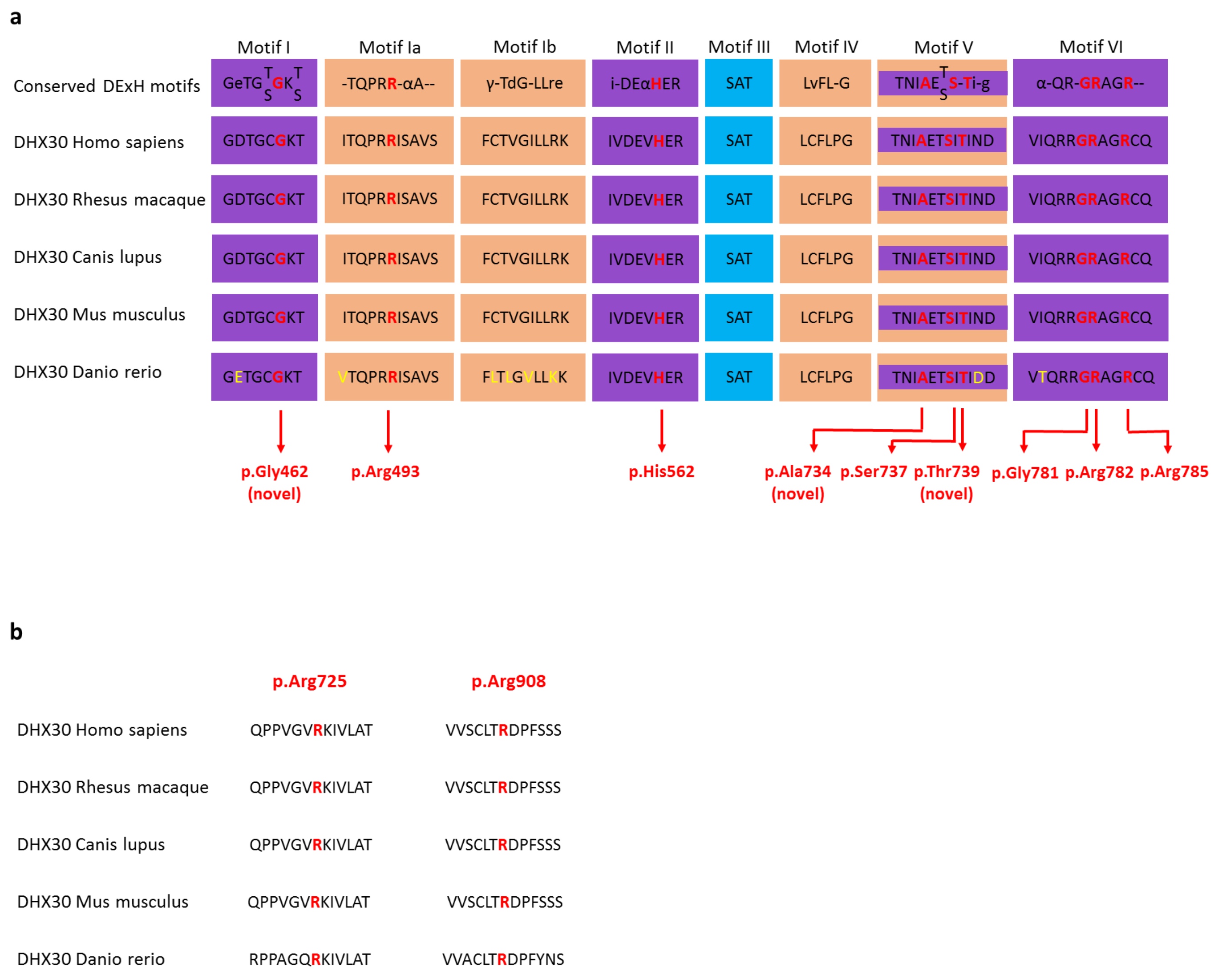


**Supplementary Figure 1. Identified missense variants affect highly conserved amino acids.** (**a**) Evolutionary conservation of the missense variants within motifs of the helicase core region. The position of the missense variants identified are shown in red. Amino acid residues affecting novel missense variants are noted in brackets. Non conserved amino acid are shown in yellow. Nucleotide-interacting motifs (I, II and VI) are shown in purple, nucleic acid-binding motifs (Ia, Ib and IV) in orange, motif V, which binds nucleic acid and interacts with nucleotides, in purple and orange, and motif III, which couples ATP hydrolysis to RNA unwinding, in blue (as previoulsy described by Lessel et al., 2017). (**b**) Evolutionary conservation of the missense variants p.(Arg725His) and p.(Arg908Gln) not located within motifs of the helicase core region. The position of the missense variants are shown in red. Note that the affected amino acids are evolutionary highly conserved from humans to zebrafish.

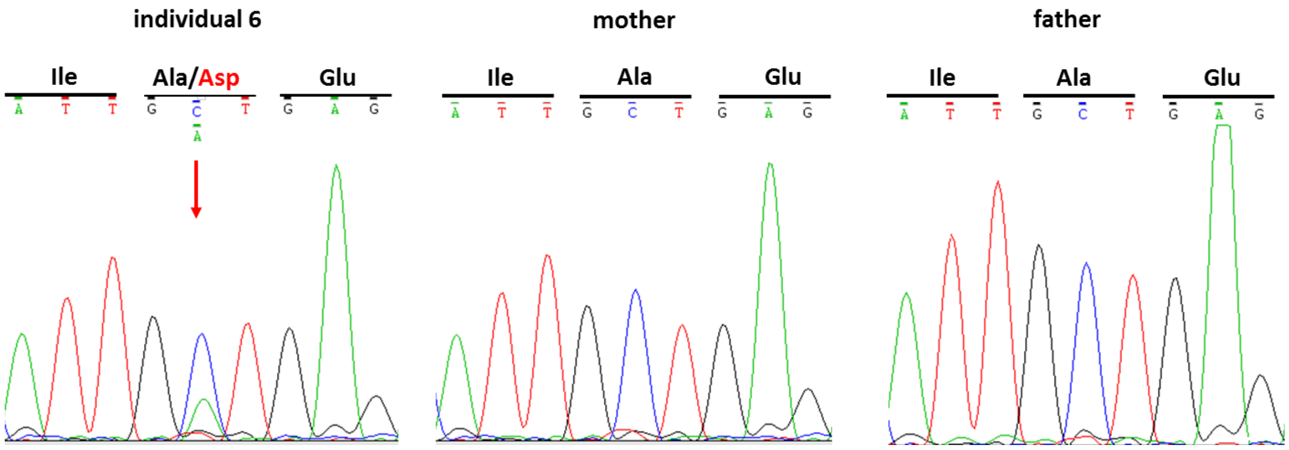


**Supplementary Figure 2. *De novo* mosaicism in individual 6.**Sanger sequence electropherograms of parts of *DHX30* after PCR amplification of genomic DNA of the affected individual 6 and her parents, confirming *de novo* mosaicism. The amino acid translation is shown in the three-letter code above the DNA sequence. The red arrow indicates the variant at c.2201C>A, p.(Ala734Asp) present only in the DNA sample of the affected individual.

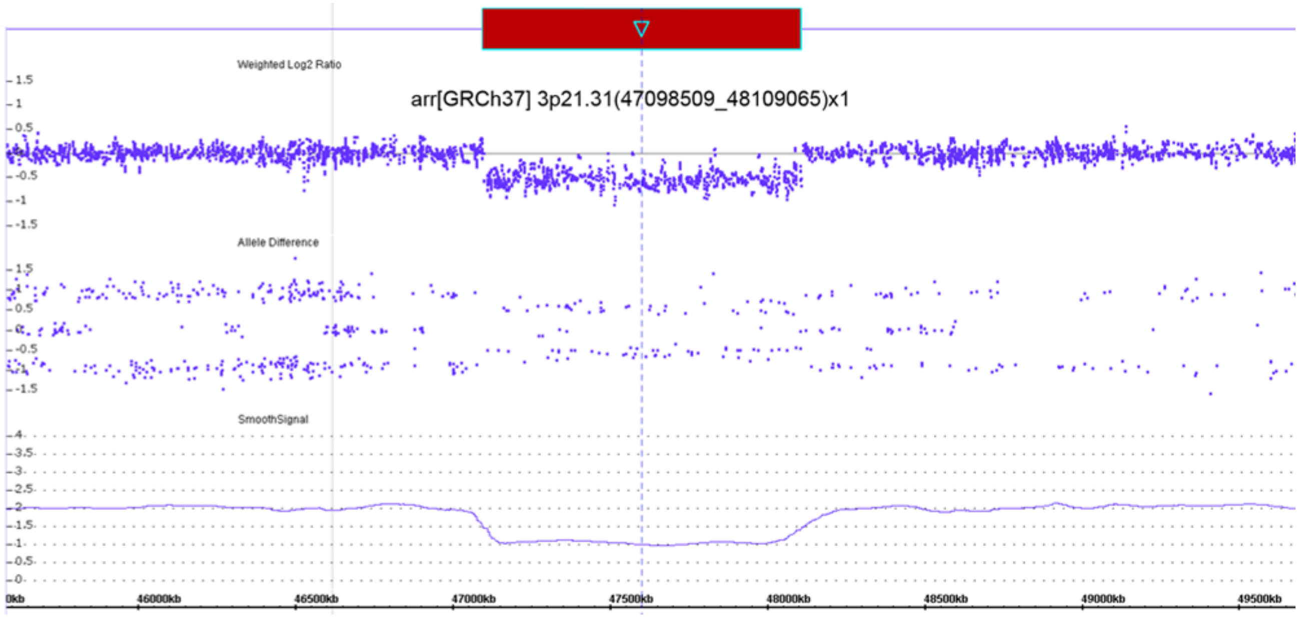


**Supplementary Figure 3. Whole gene deletion in individual 24.** Adapted from Chromosome Analysis Suite 3.3 (ChAS 3.3) showing loss of oligonucleotide probes at 3p21.31. Each dot represents one single nucleotide polymorphism that are distributed on the x-axis which shows the genomic positions.

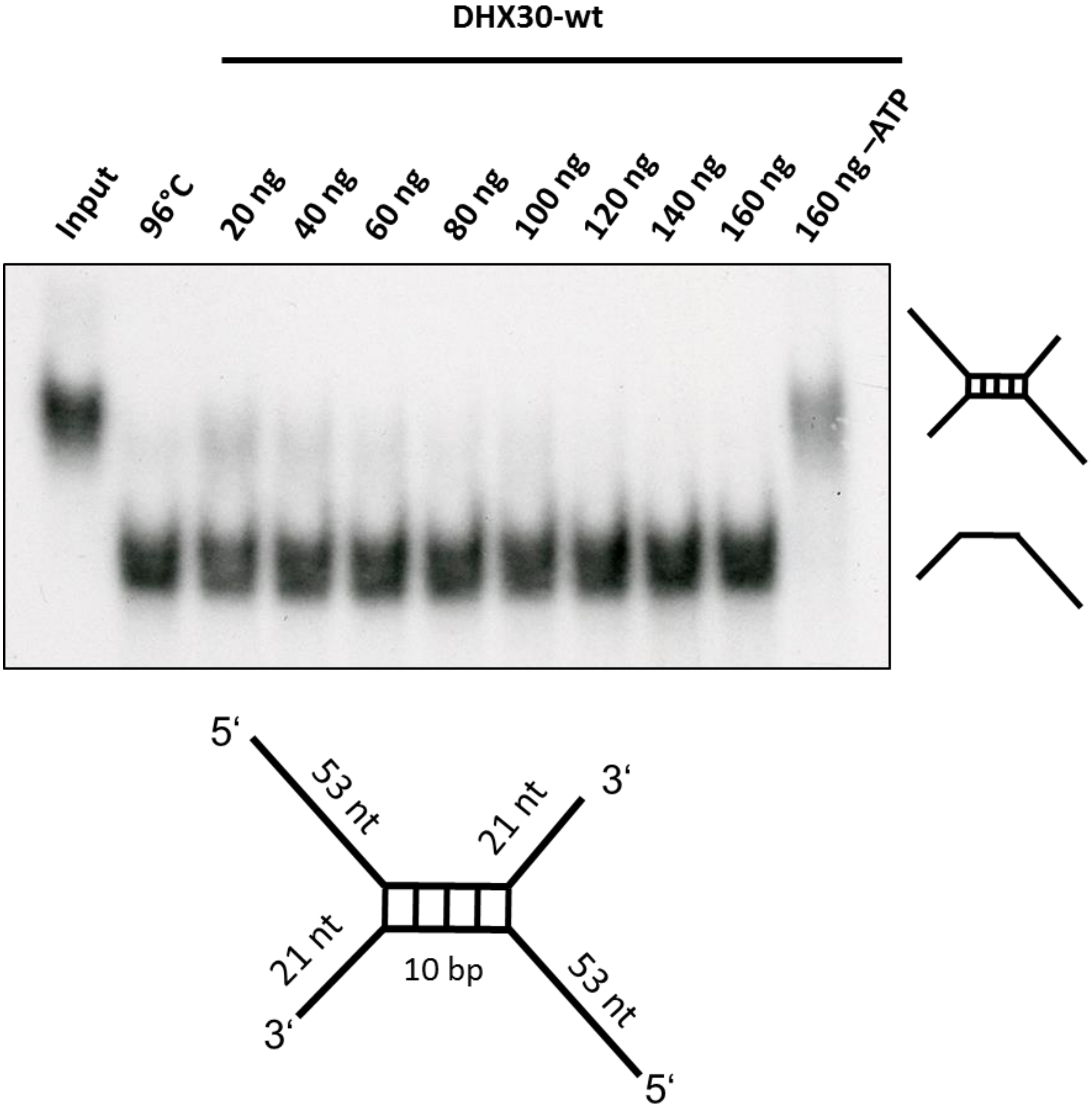


**Supplementary Figure 4. DHX30 WT acts as an ATP-dependent RNA helicase.** Top: Increasing amounts of His6-SUMO-tagged DHX30 WT protein were incubated with a 32P-labelled RNA substrate in the presence (lane 3-7) or absence (lane 8) of ATP and analyzed by native PAGE. The position of the RNA duplex and the single-stranded RNA are indicated in the first and second lane respectively. Their schematic representation is shown at the right side. Bottom: RNA duplex containing a central GC sequence flanked by single-stranded regions of 53 nucleotides at the 5’ end and 21 nucleotides at the 3’ end.

**a**

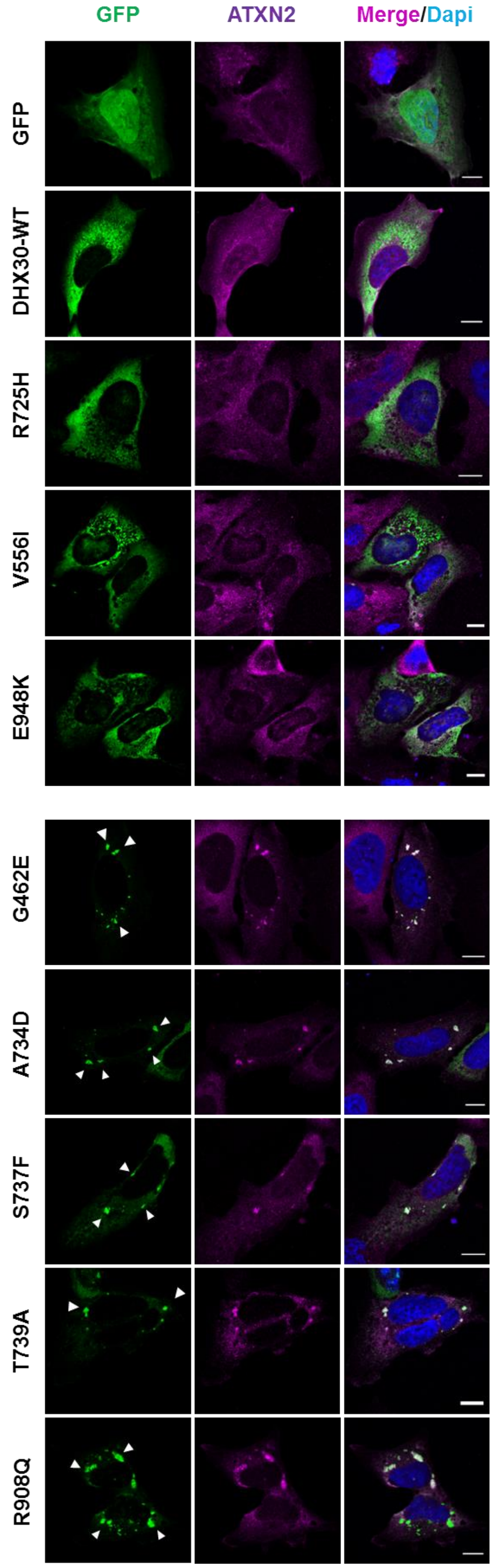
GFP
ATXN2
Merge/Dapi
GFP
DHX30-WT
R725H
V556I
E948K
G462E
A734D
S737F
T739A
R908Q

**b**

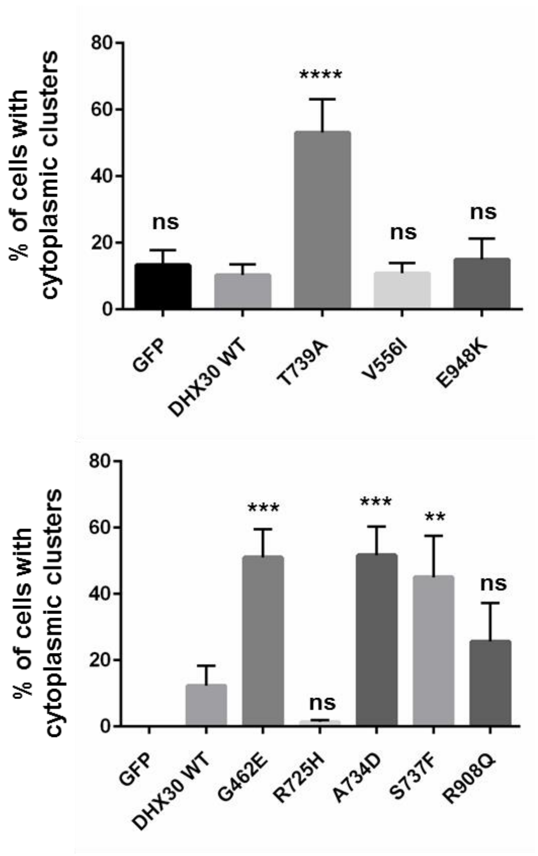
% of cells with cytoplasmic clusters
80
60
40
20
0
ns
****
ns
ns
GFP
DHX30 WT
T739A
V556I
E948K
% of cells with cytoplasmic clusters
80
60
40
20
0
***
ns
***
**
ns
GFP
DHX30 WT
G462E
R725H
A734D
S737F
R908Q

**Supplementary Figure 5. Recombinant protein variants of DHX30 induce the formation of cytoplasmic clusters.** (**a**) Immunocytochemical detection of DHX30-GFP fusion proteins (GFP, green) and endogenous ATXN2 (magenta) in transfected U2OS cells. Upper panel: wild-type DHX30-GFP preferentially resides throughout the cytoplasm and GFP accumulates in nuclei, similar to recombinant protein variants of DHX30 harboring amino acid substitutions V556I, R725H and E948K (upper panel). Lower panel: recombinant protein variants of DHX30 harboring amino acid substitutions in the helicase core region G462E, A734D, S737F and T739A induce the genesis of cytoplasmic foci containing endogenous SG-marker ATXN2 (arrowheads), Notably R908Q amino acid substitution lead to the formation of clusters co-localizing with the SG-marker ATXN2 in only 50% of transfected cells. Nuclei are identified via DAPI staining (blue). Scale bars indicate 10 µm. (**b**) Bar graph indicating the percentage of transfected cells, in which recombinant proteins induce the emergence of clusters. (**,***,****: significantly different form DHX30-WT: **$p < 0.01$; ***$p < 0.001$; ****$p < 0.0001$; $n > 100$ from 3 independent transfections; One-Way ANOVA followed by Dunnett’s multiple comparisons test).

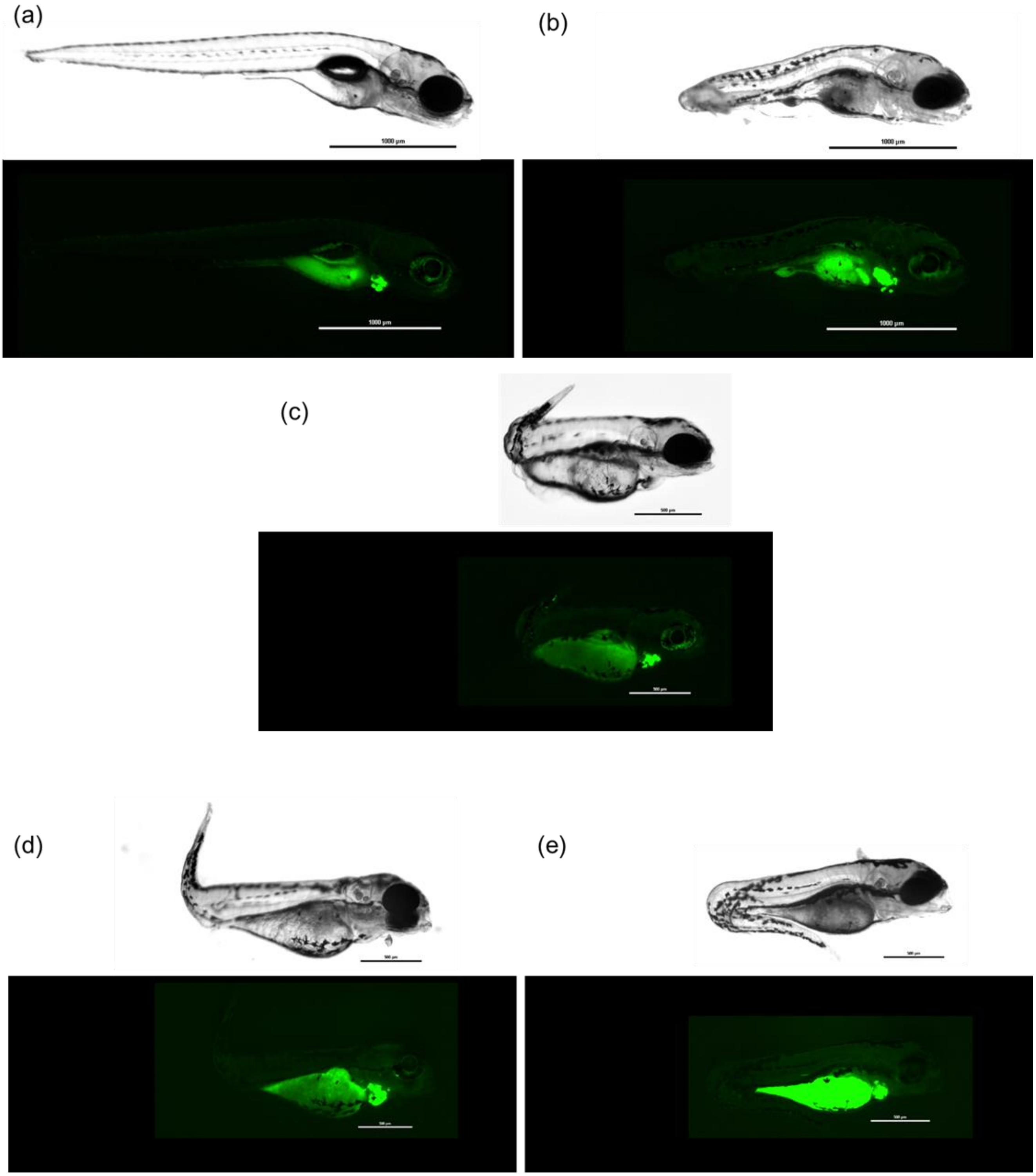


**Supplementary Figure 6. Representative images of zebrafish embryos.** (**a**) Injected with Tol2 mRNA and pTol2pA2-cmlc2:EGFP;tuba1a:DHX30 wild-type or (**b**) DHX30 harboring mutations R493H, (**c**) R725H, (**d**) R785C, or (**e**) or R908Q. Scale bars show 1000uM or 500 uM per unit. Embryos injected with wild-type DHX30 showed apparently normal development at day 7. Embryos injected with mutated DHX30 showed sign of severe developmental defects before day 7.

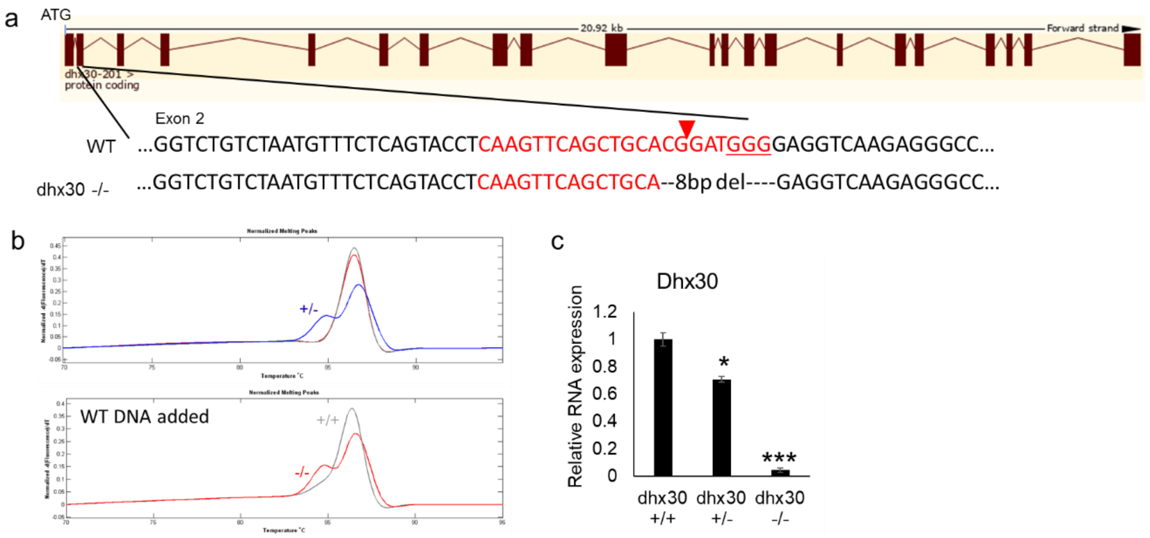


**Supplementary Figure 7. Generation of zebrafish CRISPR-Cas9-mediated dhx30 stable knockout line. (a)** Shown are genomic regions of zebrafish dhx30 targeted by CRISPR-Cas9. Red indicates gRNA-binding site with the protospacer-adjacent motif (underlined) in wild-type (WT) sequence. The mutant animals carry an 8 bp-deletions generated by CRISPR-Cas9. (**b**) Dhx30-targeted PCR products were analyzed by high-resolution melting analysis (HRM) to distinguish wild-type (+/+), heterozygous (+/-), and homozygous (-/-) animals. Two different melting peaks were shown in heterozygous PCR product (top). To distinguish wild-type and homozygous animals, wild-type DNA samples are mixed with DNA from the 'test' animals. Homozygous DNA hence become heterozygous-like, resulting in two melting peaks (bottom). (**c**) Analyses of dhx30 transcript levels in dhx30 mutant animals at 5 days post fertilization. Data are presented as means ± standard error of mean and are based on 3 replications. *, ***: significantly different from DHX30+/+ (*$p<0.05$, ***$p<0.001$; n=3; One-way ANOVA, followed by the Holm-Sidak multiple comparison test).
